# Connectome-based symptom mapping and *in silico* related gene expression in children with autism and/or attention-deficit/hyperactivity disorder

**DOI:** 10.1101/2024.12.09.24318621

**Authors:** Patricia Segura, Marco Pagani, Somer L. Bishop, Phoebe Thomson, Stanley Colcombe, Ting Xu, Zekiel Z. Factor, Emily C. Hector, So Hyun Kim, Michael V. Lombardo, Alessandro Gozzi, Xavier F. Castellanos, Catherine Lord, Michael P. Milham, Adriana Di Martino

## Abstract

Clinical, neuroimaging and genomics evidence have increasingly underscored a degree of overlap between autism and attention-deficit/hyperactivity disorder (ADHD). This study explores the specific contribution of their core symptoms to shared biology in a sample of N=166 verbal children (6-12 years) with rigorously-established primary diagnoses of either autism or ADHD (without autism). We investigated the associations between inter-individual differences in clinician-based dimensional measures of autism and ADHD symptoms and whole-brain low motion intrinsic functional connectivity (iFC). Additionally, we explored their linked gene expression patterns *in silico*. Whole-brain multivariate distance matrix regression revealed a transdiagnostic association between autism severity and iFC of two nodes: the middle frontal gyrus of the frontoparietal network and posterior cingulate cortex of the default mode network. Across children, the greater the iFC between these nodes, the more severe the autism symptoms, even after controlling for ADHD symptoms. Results from segregation analyses were consistent with primary findings, underscoring the significance of internetwork iFC interactions for autism symptom severity across diagnoses. No statistically significant brain-behavior relationships were observed for ADHD symptoms. Genetic enrichment analyses of the iFC maps associated with autism symptoms implicated genes known to: *(i)* have greater rate of variance in autism and ADHD, and *(ii)* be involved in neuron projection, suggesting shared genetic mechanisms for this specific brain-clinical phenotype. Overall, these findings underscore the relevance of transdiagnostic dimensional approaches in linking clinically-defined phenomena to shared presentations at the macroscale circuit- and genomic-levels among children with diagnoses of autism and ADHD.

## INTRODUCTION

The growing awareness of frequent comorbidities and symptom co-occurrence among neuropsychiatric conditions has motivated a shift from case-control categorical comparisons to transdiagnostic investigations of the biology underlying symptom dimensions.[1] This is particularly relevant for autism spectrum disorder (henceforth autism) and attention-deficit/hyperactivity disorder (ADHD), two common childhood-onset neurodevelopmental conditions. While autism and ADHD are conceptualized as distinct categorical diagnoses, abundant clinical evidence has documented their overlapping symptoms.[2–6] Some degree of shared genetic variance,[7–9] as well as both brain structural[10–13] and functional[14–17] atypicalities, have also been reported. Yet, despite initial insights from the clinical, neuroimaging and genomics evidence summarized below, the extent to which co-occurring symptom profiles reflect shared biological features is largely unknown.

Clinically, the co-occurrence of ADHD symptoms in children with autism has been recognized, with frequencies ranging from 28% to 80%.[18–20] More recently, the presence of symptoms qualitatively similar to autism (i.e., autistic traits) has been acknowledged in a large proportion of those with a primary ADHD diagnosis. Indeed, when examined, autistic traits have been documented in up to 32% of ADHD children.[2–6] Both patterns of symptom co-occurrence exacerbate clinical impairment, challenge accurate diagnoses, and impede individualized care.[21–23] This clinical evidence supports using transdiagnostic dimensional approaches to investigate the neurobiological underpinnings of inter-individual symptomatology differences in both symptom domains.

From a neuroimaging perspective, accumulating evidence points towards atypical widespread large-scale functional networks in both autism and ADHD.[24–28] As such, intrinsic functional connectivity (iFC) measured with resting-state functional MRI (R-fMRI) is useful to investigate shared atypical neural features. To date, only a few R-fMRI studies have directly examined associations between iFC and dimensional measures of autism and/or ADHD symptoms across individuals with categorical diagnoses of autism or ADHD in the same study.[15–17] However, findings have been inconsistent. One study did not find significant iFC associations specific to either autism or ADHD symptoms.[16] The other two studies revealed significant transdiagnostic iFC associations,[15, 17] but the relative contribution of each targeted symptom domain remains unspecified. For example, the iFC patterns predicted by parent-reported autism severity in an autism dataset were found to be associated with parent ratings of inattention in an independent ADHD sample.[17] However, the lack of autism measures in the independent ADHD dataset prevented assessing the degree to which the identified iFC pattern was uniquely related to the autism domain. As a result, the specific contribution of core autism or ADHD symptoms to the iFC atypicalities shared across children with categorical diagnoses of autism and/or ADHD remains uncertain.

At the genomic level, accumulating evidence also converges on shared genetic mechanisms across autism and ADHD.[8, 9] These highly heritable conditions[29, 30] have been found to co-aggregate in families with a parent or a child with either of these diagnoses,[31, 32] to share the same comorbidity patterns among monozygotic twins,[32, 33] and to be genetically correlated (rG=0.36).[34] For both autism and ADHD, genetic liability is thought to involve polygenic effects of common variants — most still undiscovered — and the role of rare — heritable and *de novo* — variants.[35, 36] Among those identified, many genes implicated in autism and ADHD are highly expressed in the brain and encode for proteins involved in transcription regulation, synaptic transmission, intracellular signaling, and nervous system development.[37, 38] Whether the expression pattern of such genes is linked to transdiagnostic brain connectivity associated with core symptoms of autism and/or ADHD is unknown.

To clarify the specific contribution of symptom dimensions to shared biological features across autism and ADHD, the present study aims to identify associations between inter-individual differences in core autism and ADHD symptoms, iFC, and their related gene cortical expression patterns. We examined iFC relationships with clinicians’ expert ratings of autism and ADHD symptom severity in school-aged children without intellectual disability and rigorous diagnoses of either autism or ADHD without autism. To assess the unique relative contribution of each symptom domain, all children underwent the same phenotypic protocol. Given that multiple brain networks have been implicated in autism and ADHD,[24, 25, 27, 28] we conducted a whole-brain unbiased investigation using multivariate connectome-based analyses.[39] Further, to address reproducibility concerns in the field, we assessed robustness of iFC-behavioral findings to different methodological choices in MRI processing, data collection, and behavioral measures. Finally, building on the notion that genetic variation contributes to iFC,[40, 41] we explored if genes reported to harbor variants associated with ADHD and autism[42] were significantly enriched among those expressed within symptom-related iFC maps.

## METHODS

### Sampling

Data were collected prior to the WHO COVID-19 pandemic. Children were mostly recruited from the New York metropolitan area from clinical and/or community sources aiming to reach parents with concerns related to autism and/or ADHD, regardless of prior diagnostic history. We included participants aged 6-to-12 years who were identified as ASD or as ADHD without ASD (i.e., ADHD_w/oASD_) and completed at least one structural T1-weighted (T1w) and one resting-state functional MRI (R-fMRI) scan passing quality assurance. Inclusion in the ASD group required clinician’s best estimate of ASD per the Diagnostic and Statistical Manual of Mental Disorders, 5th Edition (DSM-5)[43] with or without any psychiatric comorbidities, including ADHD, given its high prevalence in ASD.[18, 20] Inclusion in the ADHD_w/oASD_ group required clinician’s best estimate of meeting ADHD DSM-5 criteria in the absence of meeting DSM-5 criteria for ASD; other comorbid diagnoses for this group were not exclusionary. Inclusion also required a full IQ > 65; a complete list of inclusion/exclusion criteria is in Supplementary material. Over the course of the study, the enrolling and phenotyping site transferred from the NYU Grossman School of Medicine Child Study Center to the Child Mind Institute (CMI). As shown in Table S1, sample characteristics did not differ by enrollment site. The study was conducted in compliance with the Institutional Review Boards at NYU and Advarra Inc. at CMI. Written parental informed consent and verbal assent were provided for all children; children seven years or older also assented in writing.

### Clinical and socio-demographics

Phenotyping included direct child assessment and parent(s) interviews, as well as review of available parent questionnaires, teacher and/or prior child’s records. Child assessments included the Differential Ability Scales-2nd Edition (DAS-II)[44] and research-reliable administration/scoring of the Autism Diagnostic Observation Schedule-2^nd^ Edition (ADOS-2)[45] by an evaluator blind to the child’s presumptive diagnosis, prior history and records. For children treated with stimulants, assessments occurred after their discontinuation for at least 24 hours. The clinician-based parent interviews included the Kiddie-Schedule for Affective Disorders and Schizophrenia for School-Age Children–Present and Lifetime Version (KSADS)[46] and the Autism Symptom Interview for verbal children,[47] conducted by an evaluator unblind to current and past concerns/records. Whenever feasible, to further characterize the sample, the Vineland Adaptive Behavior Scales-2^nd^ Edition (VABS-II)[48] parent interview was collected along with parent questionnaires assessing symptoms of autism, ADHD, and general psychopathology using the Social Responsiveness Scale-2^nd^ Edition (SRS-2),[49] the Social Communication Questionnaire-Lifetime (SCQ-L),[50] the Strengths and Weaknesses of ADHD Symptoms and Normal Behavior (SWAN),[51] and the Child Behavior Checklist (CBCL).[52] Information about ADHD-related symptoms in school were obtained via SWAN teacher questionnaires, whenever feasible. Socioeconomic status, indexed by the Hollingshead scale,[53] and parent-reported ethnic/racial backgrounds were also collected.

### Diagnostic protocol

Best estimate clinician-based DSM-5 psychiatric diagnoses were established with a rigorous multistep team-based approach. First, following their assessments, the blind and unblind evaluators (i.e., child and parent evaluator, respectively) met to share their impressions and records to reach an initial diagnostic consensus in the primary diagnostic groups. The evaluators provided ratings of diagnostic certainty for the presence or absence of ASD (with or without ADHD) or of ADHD_w/oASD_. Then, a multidisciplinary conference reviewed all information to confirm best-estimate diagnosis of either ASD (with or without ADHD and other comorbid diagnoses) or ADHD_w/oASD_ (with or without other comorbid diagnoses), and to resolve potential diagnostic disagreements; see Supplementary material.

### MRI

Following MRI simulator training, children were invited to at least one MRI session at the NYU Center for Brain Imaging as detailed elsewhere.[54] All MRI images were collected using the same Siemens Prisma 3.0T MRI scanner with a 32-channel head coil (Siemens; Erlangen). T1w images were obtained using a magnetization prepared gradient echo sequence, followed by at least a 6.33 min R-fMRI scan (Table S2). During R-fMRI acquisition, participants were instructed to keep their eyes open, relax and remain still. Whenever feasible, children completed an additional 4.65 min R-fMRI scan. Children treated with stimulant medications discontinued them at least 24 hours prior to MRI.

### MRI data quality assurance (Q/A) and preprocessing

Structural T1w scans were visually inspected for artifacts (e.g., severe ringing, blurring, ghosting, or gross anatomical abnormalities) by one of two raters with excellent interrater reliability established on 84 study images (ICC_(3,1)_: 0.96; 95% CI: 0.94-0.97). For R-fMRI data, following visual inspection for signal dropouts/artifacts, only scans with a median framewise displacement[55] (FD) ≤0.2*mm* were analyzed. Using this cutoff, of the 194 children meeting inclusion criteria and completing at least one MRI session, 28 did not pass Q/A, and thus were excluded from analyses. No differences were seen between those excluded and included with respect to demographics, intelligence, or key symptom ratings (Table S3). For further characterization of Q/A data, we computed individual-level Euler index[56] for T1w data and the number of R-fMRI peaks with FD≥1*mm*. Overall, motion was low across the sample analyzed, with no differences by primary diagnosis (Table 1, Fig. 1c).

**Fig. 1:**
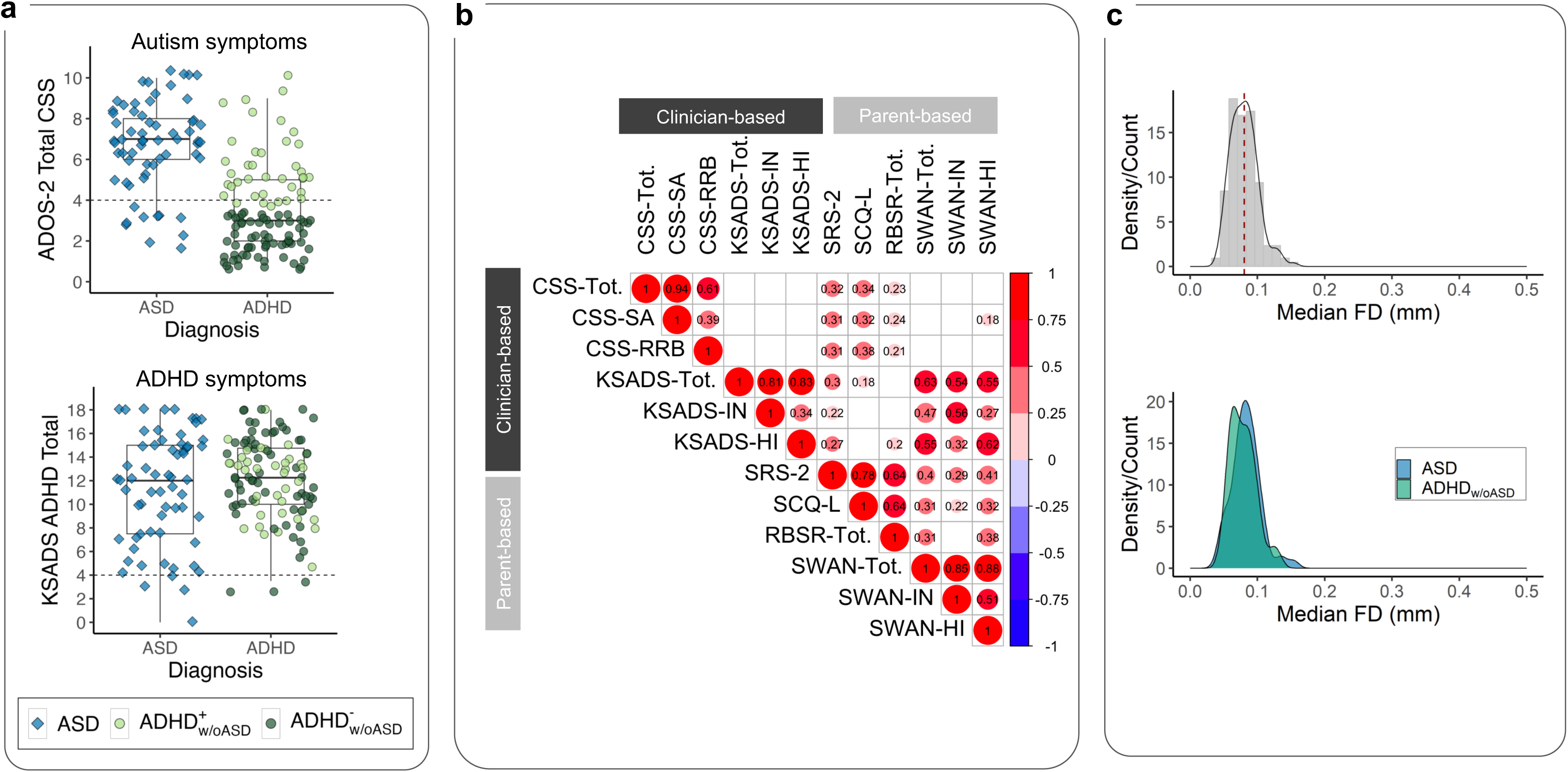
Key characteristics of the sample. **a)** We included N=166 children comprising n=63 classified with autism and n=103 classified as ADHD_w/oASD_ (blue diamonds and green circles, respectively). The box plots show the distribution of clinician-based ratings of autism and ADHD symptoms, indexed by the ADOS-2 CSS-Total (top) and KSADS total scores (bottom), respectively for the children divided by their primary diagnostic group. As expected, on average, the children in the autism group had significantly elevated CSS-Total scores. While on average, children in the ADHD_w/oASD_ group had lower CSS-Total scores than the autism group; 38 (37%) of these children were at or above the ADOS-2 CSS-Total cutoff for autism spectrum (i.e., CSS-Total=4). This subset of children, labeled ADHD_w/oASD_^AS+^, is shown as dark green circles, the remaining (ADHD_w/oASD_^AS-^) are marked as light green circles. **b)** Correlation matrix across autism and ADHD symptom severity measures based on clinician’s ratings (ADOS-2 observation and KSADS parent interview) and parent questionnaire scores (SRS-2, SCQ, and SWAN, respectively). Only correlations surviving FDR-correction *q*<0.05 are shown in the matrix, with circle size indicating the correlation magnitude. Notably, unlike autism severity based on parent responses (i.e., SCQ and SRS-2), ratings based on clinicians’ observation (i.e., ADOS-2) were not significantly related to any ADHD ratings (i.e., SWAN and KSADS). **c)** Head micromovement distribution indexed by median framewise displacement (FD) across all 166 children included in analyses (top gray density plot), and by diagnostic group (bottom plots; blue autism and green ADHD_w/oASD_). Overall head motion was low with no significant differences between diagnostic groups. Abbreviations: ADOS-2 CSS-Total, Autism Diagnostic Observation Scale-2^nd^ Edition Total Calibrated Severity scores; KSADS, Kiddie-Schedule for Affective Disorders and Schizophrenia for School-Age Children–Present and Lifetime Version; SRS-2, Social Responsiveness Scale-Second edition; SCQ-L, Social Communication Questionnaire-lifetime; SWAN, Strengths and Weaknesses of ADHD Symptoms and Normal Behavior.

**Table 1.** Characteristics of the sample combined and by primary diagnostic groups.

| Variable | Total<br>N=166 | ASD<br>n=63 | ADHD w/o ASD<sup>1</sup><br>n=103 | Statistics |  |  |  |
| --- | --- | --- | --- | --- | --- | --- | --- |
| | | | | df | W $\chi^2$ | p value | FDR-p value |
| Age, years, M (SD) | 8.9 (1.7) | 8.8 (1.9) | 8.9 (1.7) | - | 3419.0 | 0.56 | 0.67 |
| Sex assigned at birth, males #, (%) | 125 (75) | 53 (84) | 72 (70) | 1 | 3.5 | 0.06 | 0.11 |
| Ethnicity, Hispanic #, (%)<sup>a</sup> | 39 (24) | 16 (26) | 23 (22) | 1 | 0.1 | 0.75 | 0.79 |
| Race, #, (%)<sup>a,b</sup> |  |  |  |  |  |  |  |
| White | 95 (58) | 32 (52) | 63 (61) | 4 | 5.3 | 0.26 | 0.34 |
| Mixed | 26 (16) | 8 (13) | 18 (17) |  |  |  |  |
| Black/African American | 19 (12) | 9 (15) | 10 (10) |  |  |  |  |
| Asian | 14 (8) | 6 (10) | 8 (8) |  |  |  |  |
| Other | 11 (7) | 7 (11) | 4 (4) |  |  |  |  |
| Socio-economic status, class #, (%)<sup>c</sup> |  |  |  |  |  |  |  |
| Classes 1-3 | 45 (28) | 19 (32) | 26 (26) | 1 | 0.4 | 0.53 | 0.66 |
| Classes 4-5 | 116 (72) | 41 (68) | 75 (74) |  |  |  |  |
| Psychoactive Medication Status, #, (%)<sup>d</sup> |  |  |  |  |  |  |  |
| Med naïve | 117 (72) | 39 (65) | 78 (76) | 2 | 5.1 | 0.08 | 0.14 |
| Currently on medication | 34 (21) | 18 (30) | 16 (16) |  |  |  |  |
| Not Naïve but current off treatment | 13 (7) | 4 (5) | 9 (9) |  |  |  |  |
| Psychiatric Comorbidity (yes), # (%) | 108 (65) | 59 (94) | 49 (48) | 1 | 34.5 | p<0.0001 | p<0.0001 |
| DAS-II IQ Standard scores, M (SD) |  |  |  |  |  |  |  |
| Verbal IQ | 107 (16) | 105 (17) | 108 (16) | - | 3565.5 | 0.29 | 0.40 |
| Non-verbal VIQ | 104 (18) | 103 (19) | 104 (18) | - | 3294.5 | 0.87 | 0.87 |
| Full-scale IQ | 105 (17) | 103 (19) | 105 (15) | - | 3450.5 | 0.49 | 0.65 |
| ADOS-2, M (SD)<sup>e</sup> |  |  |  |  |  |  |  |
| Expressive Language score<sup>f</sup> | 7.9 (0.4) | 7.8 (0.6) | 7.9 (0.3) | - | 3604.5 | 0.03 | 0.06 |
| SA CSS | 5.1 (2.6) | 6.9 (2.1) | 4.1 (2.3) | - | 1230.5 | p<0.0001 | p<0.0001 |
| RRB CSS | 4.6 (3.2) | 6.8 (2.9) | 3.3 (2.7) | - | 1297.5 | p<0.0001 | p<0.0001 |
| Total CSS | 4.7 (2.7) | 6.7 (2.2) | 3.4 (2.1) | - | 992.5 | p<0.0001 | p<0.0001 |
| SRS-2 Parent Total T scores, M (SD) | 62 (12.2) | 70.2 (9.7) | 57 (10.8) | - | 1184.5 | p<0.0001 | p<0.0001 |
| SCQ-Lifetime Total, M (SD)<sup>g</sup> | 9.7 (7.7) | 15.5 (6.7) | 6.2 (5.9) | - | 783.5 | p<0.0001 | p<0.0001 |
| ASI Total, M (SD) | 35.8 (14.9) | 45.5 (11.3) | 30 (13.9) | - | 1195.0 | p<0.0001 | p<0.0001 |
| Vineland Adaptive Functioning-II, M |  |  |  |  |  |  |  |
| Communication | 81.9 (10.6) | 79.9 (11.2) | 83.2 (10) | - | 3600.5 | 0.04 | 0.07 |
| Socialization | 83.4 (14.4) | 76.6 (12.9) | 87.8 (13.6) | - | 4323.5 | p<0.0001 | p<0.0001 |
| Daily Living | 81.1 (10.9) | 77.4 (11) | 83.5 (10.2) | - | 4013.0 | p<0.0001 | 0.001 |
| ABC Composite scores | 80 (10.9) | 75.4 (11.7) | 83 (9.3) | - | 4195.5 | p<0.0001 | p<0.0001 |
| K-SADS ADHD severity, M (SD) |  |  |  |  |  |  |  |
| Inattentive | 6.8 (2.2) | 6.4 (2.5) | 7.1 (2) | - | 3668.0 | 0.16 | 0.24 |
| Hyperactive | 5.6 (2.4) | 5.5 (2.6) | 5.7 (2.4) | - | 3332.5 | 0.77 | 0.80 |
| Total | 12.4 (3.8) | 11.9 (4.4) | 12.8 (3.3) | - | 3541.0 | 0.32 | 0.44 |
| SWAN Parent Averaged scores, M (SD) |  |  |  |  |  |  |  |
| Inattentive | 1.1 (0.9) | 1.1 (0.9) | 1.2 (0.9) | - | 3397.5 | 0.61 | 0.69 |
| Hyperactive | 0.9 (1) | 1.1 (1) | 0.8 (1) | - | 2642.0 | 0.05 | 0.09 |
| Total | 1 (0.8) | 1.1 (0.8) | 1 (0.8) | - | 2982.0 | 0.38 | 0.52 |
| SWAN Teacher Averaged scores, M (SD)<sup>i</sup> |  |  |  |  |  |  |  |
| Inattentive | 1.1 (0.83) | 1.09 (0.723) | 1.12 (0.9) | - | 582.5 | 0.65 | 0.72 |
| Hyperactive | 0.86 (0.89) | 0.76 (0.83) | 0.92 (0.9) | - | 587.0 | 0.61 | 0.71 |
| Total | 0.98 (0.72) | 0.92 (0.65) | 1.02 (0.77) | - | 584.0 | 0.64 | 0.72 |
| CBCL T scores, M (SD)<sup>j</sup> |  |  |  |  |  |  |  |
| Internalizing | 58.5 (10.6) | 62.4 (9.3) | 56.2 (10.7) | - | 2129.0 | p<0.0001 | 0.001 |
| Externalizing | 58.8 (10.2) | 60.4 (10.6) | 57.9 (9.9) | - | 2770.5 | 0.16 | 0.25 |
| Total Problems | 62.3 (8.9) | 65.8 (8.1) | 60.2 (8.8) | - | 2059.5 | p<0.0001 | p<0.0001 |
| In-scanner motion, median FD, M (SD) |  |  |  |  |  |  |  |
| R-fMRI-1 6'20" | 0.08 (0.02) | 0.08 (0.02) | 0.08 (0.02) | - | 2784.5 | 0.13 | 0.21 |
| R-fMRI-2 4'39"<sup>k</sup> | 0.14 (0.04) | 0.1 (0) | 0.1 (0) | - | 967.0 | 0.15 | 0.23 |
| # Peaks $\geq 1$ mm, M (SD) | | | | | | | |
| R-fMRI-1 6'20" | 2.4 (6.8) | 3.4 (8.6) | 1.8 (5.3) | - | 2794.0 | 0.07 | 0.12 |
| R-fMRI-2 4'39"<sup>k</sup> | 2.8 (7.4) | 4.6 (11.2) | 2.1 (4.6) | - | 1003.5 | 0.18 | 0.24 |
| Euler index | -112 (76.6) | -117 (78.1) | -110 (75.9) | - | 3301.0 | 0.77 | 0.79 |

Diagnostic group differences (i.e., Autism Spectrum Disorder [ASD] vs Attention Deficit Hyperactivity Disorder without ASD [ADHDw/oASD]) were tested with Wilcoxon Rank sum tests for continuous variables and with Pearson Chi-square tests for categorical variables. p-values were corrected with False Discovery Rate (FDR) at $q > 0.05$ ; those remained significant after correction are shown in bold.

Preprocessing was performed using the Configurable Pipeline for the Analysis of Connectomes (Fig S1; supplementary material).[57] Briefly, preprocessing steps for T1w images included intensity non-uniformity correction, skull stripping, tissue segmentation, and spatial normalization using ANTS. For R-fMRI scans, nuisance signal regression followed realignment to anatomical images and skull-stripping and included temporal bandpass filtering (0.01-0.1 Hz), removal of Friston 24-motion parameters, and 5-component-based noise correction (aCompCor);[58] functional-to-anatomical co-registration using FSL FLIRT’s boundary-based registration, spatial smoothing (6mm full-width-at-half-maximum) followed.

### Brain connectome-wide association analyses

#### Overview

To explore brain-behavior associations in the whole-brain, we employed a multivariate voxel-wise distance matrix regression (MDMR) discovery framework.[39, 59] MDMR identifies brain regions for which full-brain connectivity patterns vary by the behavioral variables of interest. The regions identified are then examined using seed-based voxel-wise correlation analysis (SCA) to delineate the specific iFC associated with that behavioral variable. Analyses focused on total ADOS-2 Calibrated Severity Scores (CSS-Total). We focused on total scores because autism is linked to impairment in both social communication and restricted and repetitive behaviors (RRBs)[60, 61] which have been both observed in children with ADHD.[4–6, 62, 63] Follow-up analyses explored ADOS-2 Social Affect (SA) and RRB domain calibrated subscores, separately. We explored ADHD-related iFC relationships using clinician-based ADHD total severity scores derived from the KSADS ratings, with follow-up exploration of inattention and hyperactivity/impulsivity ratings (see Supplementary material, Fig S2).

#### MDMR-based discovery framework

Consistent with the previously developed MDMR approach,[39, 59] we first calculated voxel-wise iFC using Pearson correlations at the subject level. Second, for each voxel, we computed the distances between any possible pairing of participants’ correlation matrices (iFC) to yield a group-level distance matrix. Third, we calculated the distance matrix for each behavioral score across participants and modeled the associations between each behavioral distance matrix and brain-wide connectivity distance matrix at each voxel. We restricted analyses to voxels falling within a ≥25% probability FSL-based gray matter mask common to all participants (Fig S3). The MDMR model included age, sex, and median FD as nuisance covariates. To examine the unique contributions of autism or ADHD symptom severities, both ADOS-2 CSS and ADHD-KSADS ratings were included in the model. Statistical significance was assessed with pseudo *F*-statistic testing using 5,000 permutations, and was followed by Gaussian random field theory correction at voxel-level thresholds Z>3.1 and cluster level *p*<0.01. The significant regions identified from MDMR, i.e., those that revealed significant relationships between variation in symptoms ratings and iFC, were selected as seeds for subsequent whole-brain voxel-wise SCA; for each region of interest (ROI, i.e., seed), iFC was computed using the Pearson correlation between the mean time series of the ROI and those of any other voxels included in the study’s gray matter mask. Correlation coefficients were Fisher *r*-to-*z* transformed at the individual level. Then, a group-level random effect general linear model via FSL-FEAT was conducted to assess the ROI’s iFC with the behavioral variable(s) of interest, including the same covariates described above along with each participant’s mean iFC of the seed examined. We corrected for multiple comparisons using Gaussian random field theory (Z>3.1, *p*<0.01). The MDMR code is available at http://connectir.projects.nitrc.org.

### Robustness

Secondary analyses investigated the robustness of our cluster-level findings to different methodological decisions. We assessed whether similar iFC-behavior associations identified in primary analyses employing the 24-parameter model and CompCor for nuisance regression, were observable after using either of two alternative MRI denoising pipelines, or longer scan data, or parent behavioral measures. Specifically, for alternative denoising, we selected two validated approaches — global signal regression (GSR)[64] and 36-parameter nuisance regression (36P).[65] Guided by accumulating evidence that longer scans yield more reliable results,[66, 67] we also assessed the robustness of primary findings after concatenating R-fMRI data across two rest scans (6.33+4.65=10.98 min), when available (*n*=118). For alternative symptom measures, we examined commonly used parent-based ratings including the SRS-2[49] and SCQ-L,[50] for autism, and the SWAN,[51] for ADHD symptoms. Analyses were conducted at the cluster-level, as detailed in Supplementary material. Findings with statistically significant correlations surviving FDR correction across all tests were considered robust.

### Genetic decoding and enrichment

We explored whether gene expression topographically associated with the iFC map(s) revealed by our discovery MDMR/SCA approach were significantly enriched for genes found to have greater rate of variants in autism and ADHD.[42] Consistent with previous studies,[68, 69] we carried out gene expression decoding and enrichment analyses. First, using NeuroVault[70] and the Allen Institute Human Brain Gene Expression Atlas,[71] we identified genes topographically expressed in the iFC map(s) of interest, *i.e., gene decoding.* Next, using hypergeometric-based statistical testing,[72] we assessed if the decoded gene list would include, above chance, genes implicated in autism and ADHD,[42] i.e., *gene enrichment.* The gene list we tested included 1046 protein-truncating and missense genes that have been identified in the largest exome sequence, to date, of individuals with autism and/or ADHD, (Supplementary material, Table S4).[42] Finally, to explore the biological processes potentially associated with the significantly enriched genes, we performed gene ontology (GO) searches with the Gene Ontology Consortium database[73] retaining terms surviving FDR *q*<0.05 (Supplementary material).

## RESULTS

### Sample Characteristics

Table 1 and Fig. 1 summarize the characteristics of the sample which included *n*=63 children meeting criteria for ASD and *n*=103 children for ADHD_w/oASD_ with group average intelligence within the typical range. Child and parent evaluators expressed substantial certainty for these primary diagnostic assignments (8.5±1.6 and 8.7±1.6, for ASD and ADHD_w/oASD_, respectively; 10 indicates full certainty - see Supplementary material) and inter-rater agreement was excellent (ICC_(1,1)_=0.94 and 0.91 for ASD and ADHD_w/oASD_, respectively). Consistent with the clinical literature on autism and ADHD,[18, 74] psychiatric comorbidity was frequent (*n*=108, 65%; see Table S5). Notably, within the autism group, the most represented comorbidity was ADHD, alone or in combination with other diagnoses (84%, n=53). As expected, the autism group showed significantly more severe ratings across all relevant autism measures than ADHD_w/oASD_ (Table 1, Fig. 1). However, consistent with prior reports of elevated autistic traits in ADHD,[3, 5] 37% (*n*=38) of the ADHD_w/oASD_ group fell at or above the ADOS-2 CSS-Total cutoff for autism spectrum (i.e., CSS=4; Fig. 1a) despite not fully meeting DSM-5 ASD criteria per clinician’s best estimates. Consistent with reports of high prevalence of ADHD in autism,[3, 5, 6] and with the high rate of comorbid ADHD in the autism sample, the two primary diagnostic groups did not differ in ADHD symptom severity across clinician, parent, nor teacher ratings (Fig. 1a, Table 1). Of note, parent-reported autism-related scores were significantly correlated with ADHD ratings derived by clinician’s parent interviews or questionnaires. In contrast, clinician-based ADOS-2-CSS were not significantly related to those ratings (Fig. 1b).

### Autism symptom severity is associated with connectivity between nodes of the fronto-parietal and default mode networks

#### MDMR

Across children, MDMR discovery whole-brain analyses revealed statistically significant (Z>3.1, *p*<0.01) transdiagnostic iFC relationships with ADOS-2 CSS-Total in two clusters in the left hemisphere: middle frontal gyrus (MFG), and precuneus/posterior cingulate cortex (PCC), nodes of the frontoparietal (FP) and default mode (DM) networks, respectively (Fig. 2a). Secondary analyses exploring the contribution of ADOS-2 subdomain scores, CSS-SA and CSS-RRB, yielded no connectome-wide level associations surviving our rigorous statistical threshold (Z>3.1; p<0.01; Fig S4a). MDMR did not reveal statistically significant associations with ADHD symptoms indexed by total, inattentive or hyperactivity/impulsivity ratings from clinicians’ parent interviews nor parent questionnaires (Fig S5). A similar pattern of MDMR results for ADOS-2 and KSADS-ADHD total was observed after adding full-scale IQ to the models. Accordingly, the follow-up SCA analyses focused on autism symptoms.

**Fig. 2:**
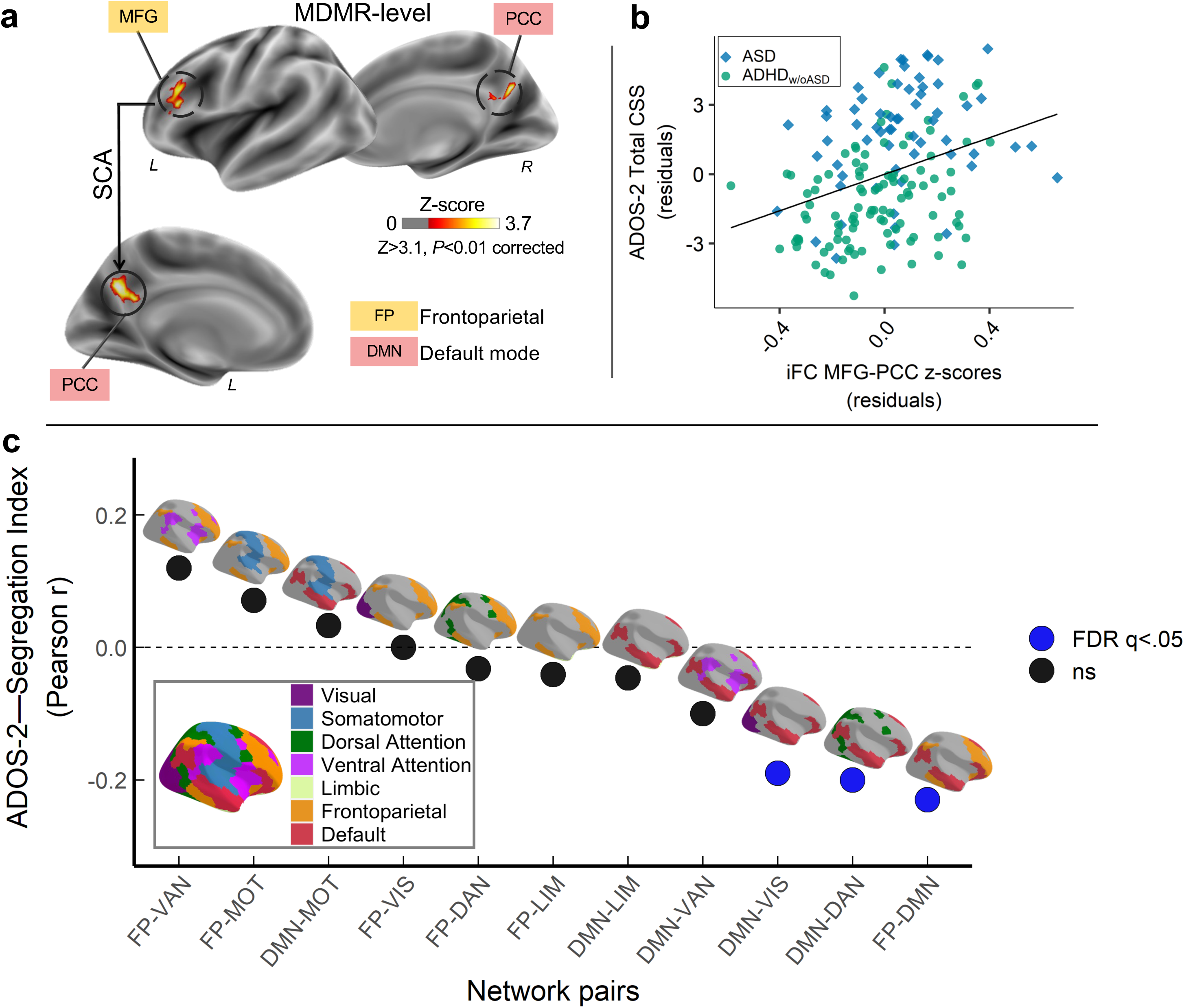
Internetwork connectivity between frontoparietal and default networks is transdiagnostically associated with autistic symptoms. **a)** Overlaid on inflated brain surface maps are the clusters with intrinsic functional connectivity (iFC) significantly associated with Autism Diagnostic Observation Scale-2^nd^ Edition Total Calibrated Severity scores (ADOS-2 CSS-Total) based on multivariate distance matrix regression (MDMR) and follow-up seed-based correlation analyses (SCA): left middle frontal gyrus (MFG: MNI152 *x*=-45, *y*=29, *z*=18) within the frontoparietal network (FP; mustard), and posterior cingulate cortex/precuneus (PCC: MNI152 *x*=-5, *y*=-59, *z*=30) within the default mode network (DMN; red). All analyses were Gaussian Random Field corrected at Z>3.1 and p<0.01. **b)** Scatter plot showing the relationship between CSS-Total and MFG iFC with PCC across children with autism spectrum disorder (ASD; blue diamonds) and Attention-Deficit/Hyperactivity Disorder without autism (ADHD_w/oASD_; green circles). Both iFC and CSS-Total are shown after regressing the same covariates used in discovery brain-behavior analyses (i.e., median FD, age, sex and KSADS ADHD total score). **c)** Correlation values (Pearson *r*) between ADOS-2 CSS-Total and segregation index (SI) between FP and DMN, as well as the SI for DMN and FP with each of the other networks defined by the 7-network Yeo-Krienen atlas (i.e., visual [VIS; purple], somatomotor [MOT; blue], dorsal attention [DAN; green], ventral attention [VAN; pink], limbic [LIM; khaki]). The SI-CSS correlations surviving FDR correction at q<0.05 are indicated as blue circles whereas black circles indicate non-significant associations. As shown, SI between DMN and FP, as well as between DMN and both DAN and VIS were significantly negatively correlated with CSS.

#### SCA

Follow-up SCA for the MDMR-identified clusters in MFG and PCC revealed a statistically significant association with CSS-Total and iFC between these nodes after controlling for KSADS ADHD total, along with the nuisance covariates listed above. Results indicated that children with higher (i.e., more severe) CSS-Totals have stronger iFC between MFG and PCC (Fig. 2b). Although MDMR analyses did not reveal significant iFC associations with CSS-SA or CSS-RRB, exploring these symptom correlations within MFG-PCC iFC indicated a statistically significant correlation with CSS-SA but not with CSS-RRB scores (*r_(164)_*=0.23, *p*=0.003; r*_(164)_*=0.08, *p*=0.290, respectively; Fig S4b).

#### ‘High’ and ‘low’ autistic traits subgroups

To further summarize findings in the context of transdiagnostic clinical presentations, we identified and compared two subgroups defined as having either ‘high’ or ‘low’ autistic traits. Specifically, regardless of their primary diagnostic status, children falling at or above the ADOS-2 autism cutoff (i.e., CSS-Total≥4) were included in the ‘high’ autistic traits subgroup, the remaining were in the ‘low’ subgroup (n=92, 55% and *n*=74, 45%, respectively). As expected, the ‘high’ autistic traits subgroup had significantly more positive MFG-PCC iFC. Both subgroups included children from the autism and ADHD_w/oASD_ primary diagnostic groups, though the ‘high’ subgroup included a statistically greater proportion of those with DSM-5 ASD diagnoses (86% of the autism sample; see Table 2). The ‘high’ subgroup had greater prevalence of males, showed elevated parent autism severity ratings and greater impairment in adaptive functioning (Table 2). Importantly, no significant subgroup differences were noted in ADHD severity measures, comorbidity rates, nor micromotion in R-fMRI data (Table 2).

#### Robustness

Cluster-level robustness analyses revealed that the association between MFG-PCC iFC and CSS-Total was robust to alternative MRI preprocessing pipelines (*r_(164)_*=0.35 and 0.34, for GSR and 36P, respectively, with FDR *q*<0.0001; Fig. 3) and to longer R-fMRI data (*r_(116)_*=0.42; FDR *q*<0.0001). Findings were not robust to the two parent-based measures of autism symptoms used as alternatives to the clinician-based ADOS-2 scores (i.e., *r_(152)_*=0.14 and 0.10, for SRS-2 and SCQ-L, respectively; FDR *q*=0.08 and *q*=0.21).

**Fig. 3:**
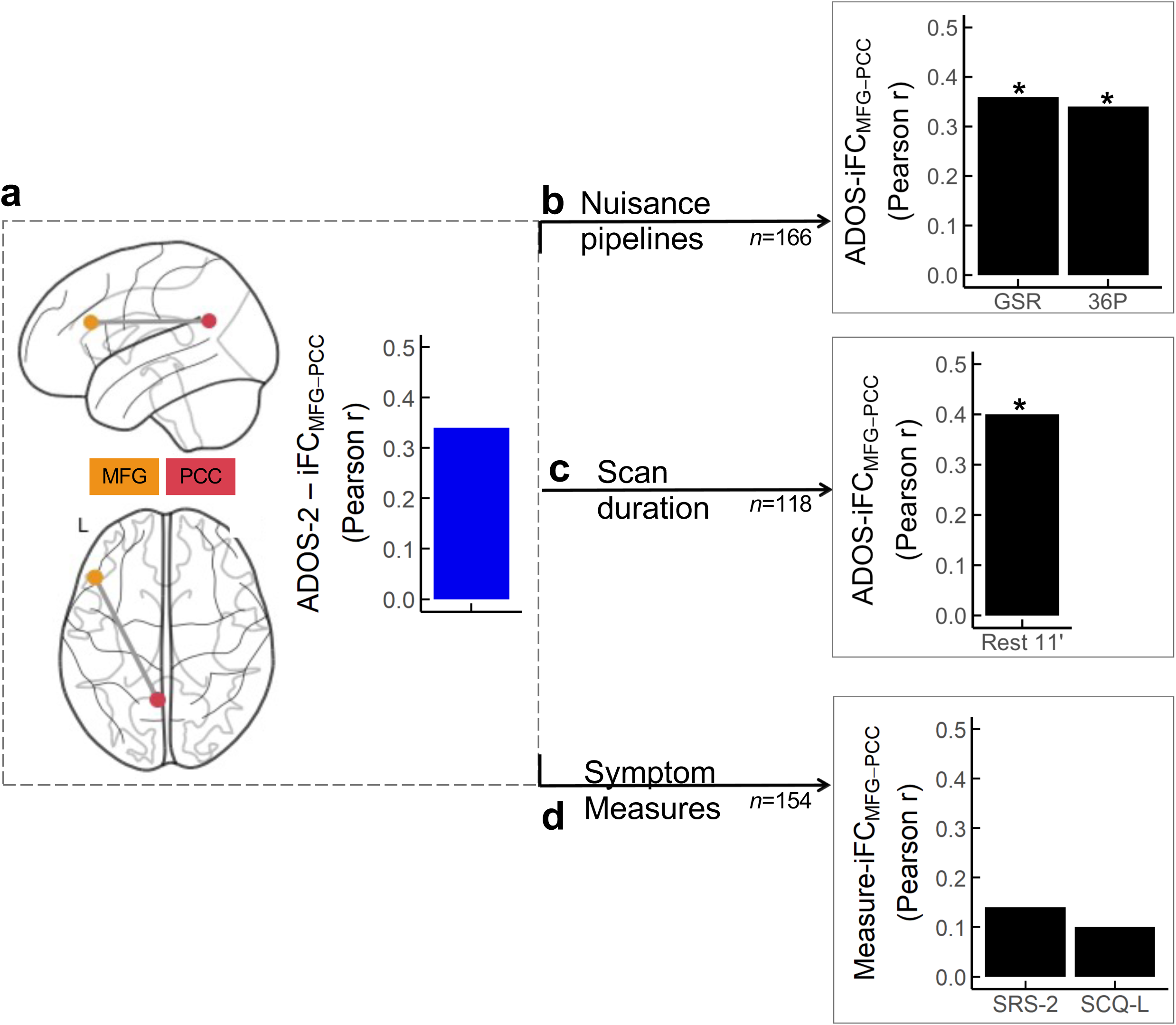
Robustness of brain-behavior primary findings to alternative MRI nuisance strategies, scan duration, and behavioral measures. **a)** Overlaid on Nilearn surface glass brains are the clusters with iFC significantly related to the Autism Diagnostic Observation Scale-second edition (ADOS-2) Total Calibrated Severity Scores (CSS-Total) based on the MDMR/SCA discovery analyses — i.e., the middle frontal gyrus (MFG) of the frontoparietal network (FP; mustard), and the posterior cingulate/precuneus (PCC) of the default mode network (DMN, red). The blue histogram illustrates the magnitude of the correlation between the MFG-PCC iFC and the ADOS-2 CSS-Total (Rest 6 minutes + discovery preprocessing pipeline, aCompCor) from the discovery analyses. The black histograms index the magnitude of the MFG-PCC correlation with ADOS-2 CSS-Total after: **b)** preprocessing using global signal regression (GSR) or 36 nuisance parameter regression (36P) in the 166 children included in the study; **c)** concatenating two quality assured resting state fMRI (R-fMRI) scans (6’20’’+4’39’’=10’59’’) in a subset of children completing both R-fMRI scans *n*=118); **d)** using parent ratings on the Social Responsiveness Scale-second edition (SRS-2) or the Social Communication Questionnaire-lifetime (SCQ-L) available in *n*=154 children. Asterisks indicate that correlations were statistically significant following FDR correction.

**Table 2.** Characteristics of the ‘high’ and ‘low’ autistic traits subgroups.

| Variable | ASD traits |  | Statistics |  |  |  |
| --- | --- | --- | --- | --- | --- | --- |
| | High<br>n=92 | Low<br>n=74 | df | F $\chi^2$ | p value | FDR-p value |
| **Primary Diagnosis, # (%)** |  |  |  |  |  |  |
| ASD | 54 (59) | 9 (12) | 1 | 35.8 | p<0.0001 | **p<0.0001** |
| ADHDw/oASD | 38 (41) | 65 (88) |  |  |  |  |
| **ADHD Presentation, # (%)** |  |  | 3 | 1.4 | 0.72 | 0.76 |
| ADHD-Combined | 39 (57) | 32 (48) |  |  |  |  |
| ADHD-Inattentive | 21 (30) | 23 (34) |  |  |  |  |
| ADHD-Hyperactive/Impulsive | 3 (4) | 3 (4) |  |  |  |  |
| ADHD Not Otherwise Specified | 6 (9) | 9 (13) |  |  |  |  |
| **Age, years, M (SD)** | 8.8 (1.8) | 9.0 (1.7) | - | 3044.0 | 0.24 | 0.32 |
| **Sex assigned at birth (males) #, (%)** | 78 (85) | 47 (64) | 1 | 8.9 | 0.003 | **0.01** |
| **Ethnicity (Hispanic) #, (%)<sup>a</sup>** | 22 (24) | 17 (23) | 1 | 0.0 | 1 | 1 |
| **Race #, (%)<sup>a,b</sup>** |  |  | 4 | 2.2 | 0.691 | 0.73 |
| White | 50 (55) | 45 (61) |  |  |  |  |
| Mixed | 16 (18) | 10 (14) |  |  |  |  |
| Black/African American | 10 (11) | 9 (12) |  |  |  |  |
| Asian | 7 (8) | 7 (9) |  |  |  |  |
| Other | 8 (9) | 3 (4) |  |  |  |  |
| **Socio-economic status class #, (%)<sup>c</sup>** |  |  | 1 | 3.0 | 0.084 | 0.14 |
| Class 1-3 | 30 (34) | 15 (21) |  |  |  |  |
| Class 4-5 | 58 (66) | 58 (79) |  |  |  |  |
| **Psychiatric Comorbidity (yes), # (%)** | 67 (73) | 41 (55) | 1 | 4.7 | 0.03 | 0.06 |
| **DAS-II IQ Standard scores, M (SD)** |  |  |  |  |  |  |
| Verbal IQ | 105 (17) | 109 (14) | - | 2852.0 | 0.07 | 0.13 |
| Non-verbal VIQ | 103 (19) | 105 (15) | - | 2954.0 | 0.14 | 0.21 |
| Full-scale IQ | 103 (19) | 106 (13) | - | 2983.0 | 0.17 | 0.23 |
| **SRS-2 Parent Total T scores, M (SD)** | 64.9 (11) | 58.4 (12.7) | - | 4509.5 | p<0.0001 | **0.001** |
| **SCQ-Lifetime Total scores, M (SD)<sup>d</sup>** | 11.1 (7.3) | 7.9 (7.8) | - | 3783.0 | 0.002 | **0.01** |
| **ASI total scores, M (SD)** | 39.9 (13.0) | 30.9 (15.7) | - | 4451.0 | p<0.0001 | **p<0.001** |
| **Vineland Adaptive Functioning-II, M** |  |  |  |  |  |  |
| Communication | 81.0 (11.3) | 83.1 (9.6) | - |  |  |  |
| Socialization | 81.1 (13.8) | 86.3 (14.6) | - | 2421.5 | 0.02 | **0.04** |
| Daily Living | 79.1 (10.1) | 83.6 (11.4) | - | 2410.0 | 0.013 | **0.03** |
| ABC Composite scores | 77.9 (11.3) | 82.6 (10) | - | 2330.5 | 0.008 | **0.02** |
| **K-SADS ADHD severity, M (SD)** |  |  |  |  |  |  |
| Inattentive | 6.46 (2.31) | 6.68 (2.43) | - | 3128.5 | 0.37 | 0.44 |
| Hyperactive | 5.15 (2.39) | 5.43 (2.66) | - | 3175.0 | 0.46 | 0.51 |
| Total | 11.61 (3.96) | 12.11 (4.08) | - | 3122.5 | 0.36 | 0.44 |
| **SWAN Parent Averaged scores, M (SD)** |  |  |  |  |  |  |
| Inattentive | 1.23 (0.89) | 1.02 (0.95) | - | 3849.0 | 0.15 | 0.21 |
| Hyperactive | 1.06 (0.9) | 0.79 (1.04) | - | 3850.0 | 0.15 | 0.21 |
| Total | 1.15 (0.77) | 0.91 (0.85) | - | 4006.0 | 0.05 | 0.10 |
| **CBCL T scores, M (SD)<sup>f</sup>** |  |  |  |  |  |  |
| Internalizing | 59.22 (10.09) | 57.68 (11.19) | - | 3631.0 | 0.39 | 0.44 |
| Externalizing | 59.38 (10.21) | 58.19 (10.25) | - | 3658.0 | 0.34 | 0.43 |
| Total Problems | 63.44 (8.68) | 60.93 (9.08) | - | 3934.0 | 0.06 | 0.12 |
| **In-scanner motion, median FD, M (SD)** | 0.08 (0.02) | 0.08 (0.02) | - | 3784.5 | 0.22 | 0.28 |
| **MFG-PCC IFC, M (SD)** | 0.13 (0.22) | 0.001 (0.23) | - | 4429.0 | 0.001 | **0.003** |

### Reduced segregation between DMN and the FP, dorsal attention and visual networks

Given that the iFC association with CSS-Total involved regions of the FP and DMN, we investigated if these brain-behavior patterns extended more broadly to FP and DMN internetwork patterns. To this end, we computed the segregation index (SI)[75] — i.e., the relative strength of within-network to between-network iFC — of each DMN and FP networks with the other functional networks defined by Yeo-Krienen[76] (see Supplementary material). Consistent with primary findings, we found significant negative associations between SI and CSS-Total such that more severe autism symptoms were associated with lower SI (higher internetwork iFC) between DMN and the FP, visual and dorsal attention networks (FDR *q*<0.05), but not with other network pairs (Fig. 2c).

### Transdiagnostic *in silico* gene expression

Given that primary SCA analyses revealed a transdiagnostic association between autism symptom severity and iFC of the MFG, we first ran gene decoding in the MFG-iFC map. This analysis identified *n*=1,519 genes topographically expressed in the MFG-iFC map (Fig. 4; gene list in GitHub). We then tested for genetic enrichment among rare genetic variants (protein truncating and missense) previously shown to have greater rates in autism and ADHD relative to controls.[42] Analyses identified significant enrichment for *n*=107 of these genes (OR=1.6, *p*=0.0002; Supplementary material, Fig. 4, Table S6). Confirming specificity of these findings, control enrichment analyses conducted using an alternative topographic gene expression map, selected to not spatially overlap with the MFG-iFC map, did not yield any significant findings (see Supplementary material). To explore the biological processes related to the 107 enriched genes, we leveraged the Gene Ontology (GO) annotation database.[73] Analyses yielded significant (FDR *q*<0.05) associations with *n*=78 GO terms (Table S7); among them most significant terms indexed neurodevelopmental morphogenesis processes involved in axonogenesis (Supplementary Material, Fig. 4b, Fig S6).

**Fig. 4:**
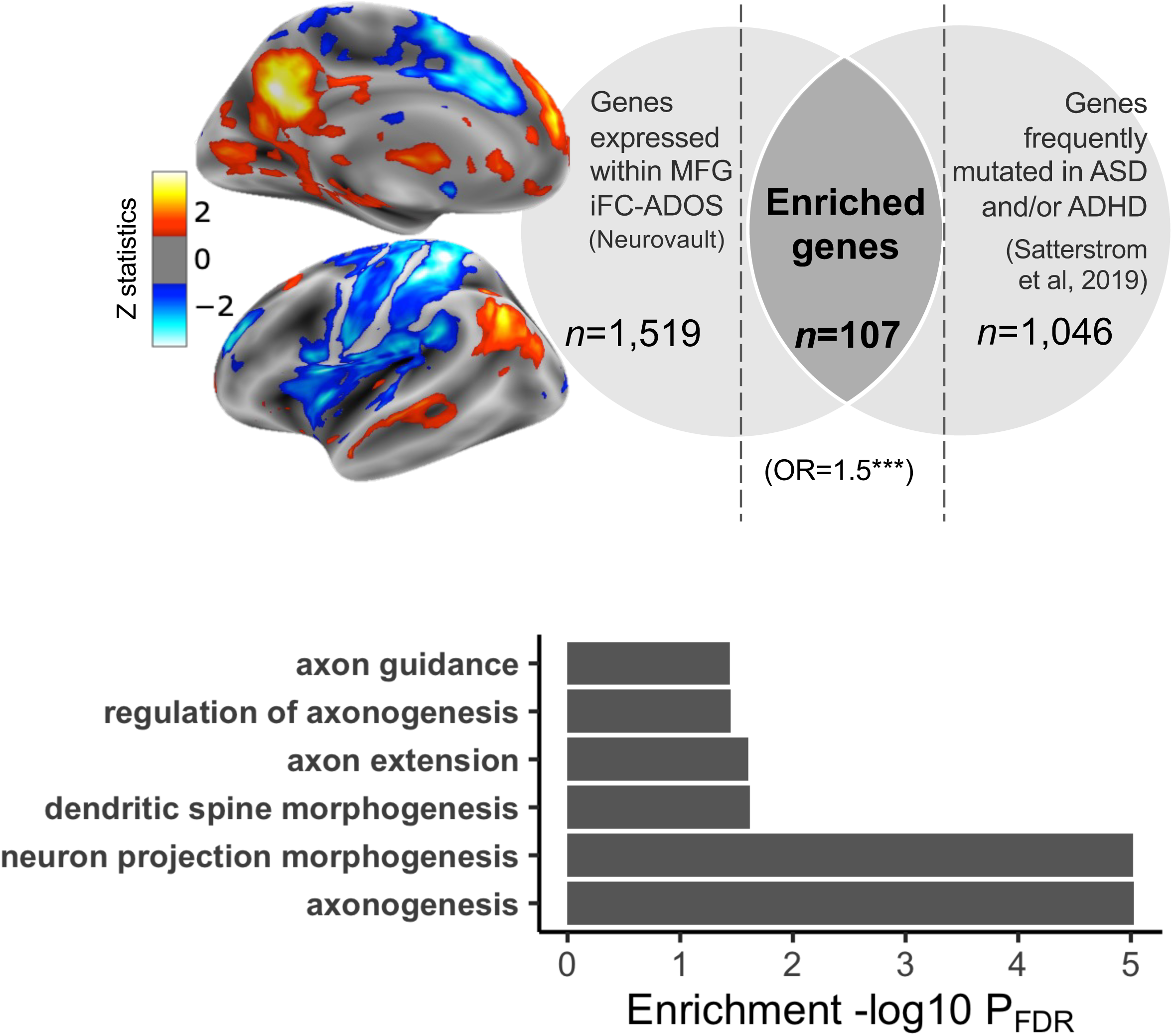
Transdiagnostic transcriptomic signature of iFC maps associated with autism symptom severity. **a.** We performed gene decoding analyses on the iFC statistical map of the middle frontal gyrus (MFG) seed associated with the Autism Diagnostic Observation Scale-2^nd^ Edition (ADOS-2) Total Calibrated Severity Scores (CSS-Total). The Z statistic MFG iFC map is overlaid on inflated surface maps. Gene decoding identified N=1,519 genes as having a spatial expression pattern significantly similar to the MFG iFC map. They are shown in the left light gray area of the Venn diagram. The right light gray area of the Venn diagram shows the number of genes (N=1,046 reported to be dysregulated in autism and/or ADHD by Satterstrom et al. 2019.[42] The number of genes resulting from gene enrichment analysis is depicted in the dark gray area of the Venn diagram as the overlap between these genes and those decoded. **b.** Specific biological GO terms included under the neuron morphogenesis term class associated with genes linked to the MFG iFC map, and related statistics (see also Fig S6 and Table S7).

## DISCUSSION

Recognizing the frequent co-occurrence of autism- and ADHD-related symptoms,[2–6] we investigated their specific contribution to iFC across school-aged verbal children with rigorously established DSM-5 diagnoses of ASD or of ADHD_w/oASD_. Whole-brain voxel-wise multivariate analyses of low motion data revealed transdiagnostic brain-behavior relationships specific to autism symptom severity, even after accounting for ADHD symptoms. Across children, inter-individual variability in symptom severity, indexed by gold-standard clinicians’ observation ratings (i.e., ADOS-2), was associated with inter-individual variability in iFC between nodes of the DMN and FP networks. Complementing results from network segregation analyses showed that autism symptom severity was inversely related to the segregation of the DMN with the FP, dorsal attention, and visual networks. *In silico* analyses revealed that gene expression within the iFC map associated with autism symptoms was significantly enriched for genes previously shown to have a higher rate of variants in individuals with autism and ADHD diagnoses without intellectual disability. Overall, our results point towards transdiagnostically shared biological underpinnings of autism symptom severity that may inform biomarker research, as well as models of vulnerability, for these neurodevelopmental conditions.

The present study underscores the role of internetwork iFC for autism-related behaviors across diagnoses. Autism-related increases in internetwork iFC have been reported in both traditional case-control comparisons[77–85] and dimensional designs within autistic samples.[77–81, 86] Our findings not only support their role among children with autism, but also elucidate their specific contribution to autism symptoms among children with ADHD_w/oASD_. Although R-fMRI studies of samples combining autism and ADHD are increasing,[87] only three studies have examined *a priori* dimensional associations of inter-individual differences in iFC with autism symptoms.[15–17] Two of them identified autism symptom associations with internetwork iFC.[15, 17] However, the autism-related iFC patterns were also associated with ADHD, thus neither study established specificity to autistic symptoms. In contrast, our phenotyping strategy of meticulously assessing both autism and ADHD symptoms in all participants allowed us to examine their relative contribution to iFC.

Additionally, prior MRI studies have primarily used parent questionnaires to index symptom severity. Although valid, these measures are known to covary with more general ratings of psychopathology (i.e., not specific to autism),[88] a finding also observed in our data. Thus, unlike prior efforts, to index variation more specific to autism we leveraged expert-based observational measures of autism severity (i.e., ADOS-2) obtained blindly to diagnostic concerns across all children.[88] Relatedly, brain-behavioral associations were weaker and statistically non-significant for parent-based ratings. In contrast, internetwork connectivity associations with ADOS-2 scores were robust to different R-fMRI analytical strategies. These findings underscore that the behavioral measures employed for brain association analyses matter, as much as the MRI metrics examined. This is in line with recent work highlighting that selecting clinical measures with high test-retest reliability increases the ability to detect relationships with brain functional metrics.[89]

Our internetwork connectivity findings involving the DMN shed new light on the role of DMN atypical connectivity previously reported in studies of either autism or ADHD.[87] By focusing on symptom domains rather than diagnostic status, our results underscore that the DMN’s interaction with other functional networks is relevant to autism symptom severity across both conditions. In typical children, internetwork iFC decreases significantly during childhood.[28, 83] These maturational processes are thought to support greater functional network segregation/specialization. Accordingly, mechanisms involved in functional network maturation may play a significant role in the emergence of autistic symptoms in children with diagnoses of autism, and, at least a subset of those with diagnosis of ADHD. The present work primarily focused on total scores of autism severity given that autism is linked to impairment in both social communication and restricted and repetitive behaviors; it was however, limited in its ability to differentiate the contribution of specific symptom subdomains. Future larger studies would benefit from identifying and leveraging finer grained autism symptom metrics to clarify whether reduced segregation of the DMN indexes a general vulnerability to autism severity or whether it has a specific role for symptom subdomains.

To our knowledge this is the first study examining, albeit *in silico*, gene expression associations with iFC in the context of brain-behavior dimensional relationships in a transdiagnostic sample including children with autism and/or ADHD. We leveraged a gene list derived from the largest exome sequencing study of autism and ADHD including protein truncating and missense genetic variants[42]. Analyses revealed that genes with a greater burden of pathogenic variants in these conditions are topographically associated with iFC patterns relevant to autism symptom severity across diagnostic groups. While this finding is consistent with models suggesting that shared genetic factors underlie disorder overlaps,[90–92] it also extends this line of work one step further by revealing that aspects of shared genetic variance among these conditions may mark vulnerability specifically for autism symptom severity rather than categorical diagnoses. This provides a more specific framework to explore previously suggested pleiotropic effects. Notably, gene ontology analyses involved terms associated to neuron morphogenesis including axonogenesis, dendritic spine morphogenesis, that have previously implicated in autism and ADHD.[93, 94] Such *in silico* associations provide hypotheses for future large-scale genome-wide association studies that can investigate shared and distinct genetic vulnerabilities of both rare and common variants.

Beyond providing biological insights, our systematic administration of clinician-based autism assessments in all children, regardless of their diagnostic concerns, detected elevated autistic traits in a substantial group of children identified as ADHD_w/oASD_. This is consistent with an emerging clinical literature recognizing that autism symptoms can be expressed in varying degrees of severity across individuals — both within and beyond diagnostic boundaries.[2–6] Recognizing children with ADHD and co-occurring autistic traits is clinically relevant. Consistent with prior work,[95] we found greater impairment in adaptive functioning in the ‘high’ autistic traits subgroup, regardless of diagnostic status (autism, ADHD_w/oASD_). Taken together, the present work highlights the importance of accounting for autistic traits when attempting to explain both the clinical and biological variance in ADHD samples.

Finally, we note that we did not observe significant connectome-wide associations specific to ADHD parent-reported severity indices, even after controlling for autism severity. Given the absence of standardized ADHD observational measures, clinician ratings were necessarily based on semi-structured parent interviews. Additionally, the relatively restricted (albeit in the clinical range) distribution of ADHD severity in the sample, may have limited the ability to detect brain-behavior associations. These observations must be placed in the context of our sample size, which, although considerably larger than the median of *N*∼23 in the literature,[96] is nonetheless only moderately large. At the same time, our findings underscore that observational behavioral measures of ADHD are needed for brain behavior associations along with a finer characterization of ADHD presentations.

Our results should be interpreted in light of limitations. First, reflecting the known preponderance of boys with these conditions, most participants were male. Thus, although we found greater prevalence of males in the ‘high’ autistic traits subgroup, this observation needs to be replicated in larger female samples. Second, as already mentioned,[96] our sample size is only moderately large and thus future efforts with better powered samples are needed. Larger transdiagnostic rigorously characterized samples will allow studies to represent, and thus assess the role of, a greater range of demographics, cognitive, as well as autism and ADHD presentations that were not assessed here.

In sum, the findings from this study enhance our understanding of the significance of internetwork iFC in autism symptom severity in children with autism and/or ADHD. They also point towards shared molecular pathways involving morphogenetic developmental processes linked to neuron projection and regulation. This work underscores the promise of exploring symptom dimensions across diagnostic categories to identify children with linked clinical and biological phenotypes and their underlying vulnerabilities. It also highlights the importance of utilizing more objective markers of behavioral phenotypes in these efforts. To realize the full potential of the framework presented in this study, future work needs to extend it to multiple dimensions across diagnoses as in ongoing efforts.[97] To fully accomplish this goal, collaborative transdiagnostic efforts using harmonized phenotyping and neuroimaging protocols are needed to gather sufficiently large samples. Such efforts will facilitate the assessment of diagnostic-specific dimensional relationships that may accompany diagnostically-shared brain-behavior associations, as shown in prior transdiagnostic studies,[98, 99] as well as enable brain-behavior subtyping to account for ADHD/ASD heterogeneity.[100]

## Data Availability

Data referred to in the manuscript are available in the National Institute of Mental Health Data Archive (NDA).

## ACKNOWLEDGEMENTS

We are deeply grateful to the children and their families participating in the study, as well as the clinical and research staff members at the Child Mind Institute and at the Child Study Center at NYU Langone Health, who supported data collection and curation. We would like to thank Steven Giavasis and the CPAC team for providing technical support for setting up the MRI preprocessing pipelines in CPAC, and to Jessica Cloud for her assistance in some aspects of MRI data quality assurance. We would also like to acknowledge the Neurovalt’s development team and the Allen Institute for their open science initiatives that enabled our *in silico* gene expression analyses. This work was partially funded by NIH R01MH105506 and R01MH115363 to ADM, R01MH091864 and R01MH120482 to MPM; the Korean Ministry of Science and ICT of the National Research Foundation RS-2023-00265410 to SHK, as well as generous gifts to the Child Mind Institute from the Dr. John and Consuela Phelan (to ADM), and from the Phyllis Green and Randolph Cowen Foundation (to MPM).

## CONFLICT OF INTEREST

Drs. Lord and Bishop receive royalties for the sale of diagnostic instruments they have co-authored (ADOS-2 and/or SCQ); profits generated from any of their own research or clinical activities are donated to charity. Dr. Di Martino is coauthor of the Italian version of the Social Responsiveness Scale — child version distributed in Italy by Organizzazioni Speciali, Italy. All other co authors report no financial interests or potential conflicts of interest.

## Notes

### Author Declarations

The study was approved by and conducted in compliance with the Institutional Review Boards of the NYU Langone Health and Advarra Inc.

