## Supplementary material for "Connectome-based symptom mapping and *in silico* related gene expression in children with autism and/or attention-deficit/hyperactivity disorder"

**Supplementary methods and results**

1. Inclusion/exclusion criteria for primary analyses
2. Diagnostic certainty
3. MRI session
4. MRI data preprocessing
5. KSADS-ADHD symptom severity
6. Gene expression decoding and enrichment
7. Segregation index
8. Control genetic decoding/enrichment analyses
9. Gene ontology analysis

**Supplementary figures**

**References**

### SUPPLEMENTARY METHODS AND RESULTS

#### 1. Inclusion/exclusion criteria for primary analyses (discovery sample)

The following were the inclusion criteria for the present study: 1) clinician's best-estimate DSM-5 diagnosis of either ASD (with or without other psychiatric comorbidity) or of ADHD without ASD (i.e., $ADHD_{w/oASD}$ ) — any presentation allowed (i.e., predominantly inattentive, combined, hyperactive impulsive); 2) completing at least one structural T1-weighted (T1w) and one resting state functional MRI (R-fMRI) scan passing our quality assurance protocols as described in the main text; 3) estimated full-scale IQ ≥ 65 — although intellectual disability was not exclusionary, ability to follow verbal instructions needed to complete the MRI scan was required; 4) English speaking. Notably, comorbid DSM-5 diagnoses were not exclusionary except for psychotic, bipolar and post-traumatic stress disorders. Other exclusion criteria for all children — i.e., regardless of ASD or $ADHD_{w/oASD}$ diagnosis — were: 1) currently active suicidal ideations, 2) history of any medical illness requiring chronic treatments, such as intrathecal chemotherapy, focal cranial irradiation, or epilepsy; 3) history of traumatic brain injury, and/or 4) contraindication for MRI scanning; 5) premature birth defined as prior to 32 weeks estimated gestational age or birth weight<1500g; 6) use of antipsychotics within six months prior to study enrollment; 7) reported history of genetic conditions known to be associated with ASD, such as Fragile X Syndrome, Tuberous Sclerosis, and Rett's Syndrome; 8) absence of signed consent by the parent or legal guardian; 9) children who did not assent regardless of parental permission; 10) children aged less than 5.5 years or greater than 12 years at enrollment. Participants treated with stimulant medications were asked to discontinue them 24 hours prior to testing and the MRI session(s). To optimize the clinical transdiagnostic sample size, this study did not include typically developing children. The sample examined in the present analyses partially overlaps with that of two prior published articles, i.e. *n*=109 (66%) and *n*=159 (96%) were included in Guttentag et al., [1] in Simhal et al.,[2] respectively.

#### 2. Diagnostic certainty

As detailed in the main text, at least two examiners were involved in the diagnostic assessment. They included a child evaluator "blind" to presumptive diagnosis and parent concerns/questionnaires, and an "unblind" parent interviewer who had access to information about presumptive diagnosis and parent-reported concerns. Following their assessments, and scoring, the "blind" and the "unblind" evaluators met to share their notes, scores and impressions, as well as to review available records to reach an initial diagnostic impression. During this process, they completed a diagnostic certainty form to quantify their best-estimate diagnosis of ASD and of $ADHD_{w/oASD}$ as also described in prior work.[1] Following the two evaluators' consensus, certainty ratings were mapped on a 1-to-10-point scale for ASD and for $ADHD_{w/oASD}$, separately. Ratings from 6 to 10 indexed the evaluator's degree of certainty of meeting criteria (for either ASD or $ADHD_{w/oASD}$), with 10 indicating highest certainty of meeting the criteria. On the other end, ratings from 1 to 5 indexed the clinicians' certainty of not meeting diagnosis of ASD or of $ADHD_{w/oASD}$, with 1 indicating highest certainty of that diagnosis' exclusion. As a note, given our diagnostic group definition, when ASD was present, it was considered the primary diagnostic designation, regardless of whether ADHD was also present or not. Across examiners, diagnostic certainty group average ratings were high for both the ASD (8.5 ± 1.6), and the $ADHD_{w/oASD}$ groups (8.7 ± 1.6). Inter-rater agreement for diagnostic certainty was excellent ($ICC_{(1,1)}$: 0.94, 95% CI = 0.93 – 0.96 for ASD and 0.91, 95% CI = 0.88 – 0.93 for $ADHD_{w/o\ ASD}$). A multidisciplinary case conference (e.g., psychiatrists, psychologists, social worker, pre-doctoral clinical psychology fellows) reviewed all information to confirm a best-estimate diagnostic consensus of either ASD (with or without comorbidities — e.g., ADHD, anxiety disorders) or $ADHD_{w/oASD}$ (with or without any other comorbidity), as well as to resolve potential disagreements, as applicable. For most children (*n*=144, 87%; ASD *n*=56, $ADHD_{w/oASD}$ *n*=88) the initial diagnostic impression of the child ("blind") and parent ("unblind") evaluators were in agreement, and, in turn, confirmed at the multidisciplinary conference. For all, but one of the remaining 22 (13%) children, the initial diagnostic impression of the two evaluators differed and a consensus was reached at the multidisciplinary conference (ASD *n*=10, $ADHD_{w/oASD}$ *n*=12). For the remaining one, the final diagnosis of ASD differed from the initial evaluators' diagnostic impression.

**3. MRI session**

As described elsewhere,[2] participants who successfully completed a full MRI simulator training were invited to an MRI session. Children unable to complete at least one good quality structural T1-weighted (T1w) and one resting state functional MRI (R-fMRI) scans passing quality control in the first session, were invited back for 'make-up' session(s), whenever possible. In the present study, MRI data from *n*=21 (13% of the total N=166 sample) were obtained at a 'make-up' MRI session.

**4. MRI data preprocessing**

All MRI data preprocessing pipelines were performed using the Configurable Pipeline for the Analysis of Connectomes (C-PAC, http://fcp-indi.github.com/C-PAC/).[3] C-PAC is an open-source software that integrates features from AFNI, FSL, ANTS and FreeSurfer to build an optimized pipeline using a Nipype python library. CPAC version 1.7.0 was used for all analyses with the exception of those focusing on 36 parameter nuisance regression, implemented with a subsequent version, CPAC version 1.8.4. Below, we detail the preprocessing steps used for the study analyses.

As shown in Figure S1a, for all analyses, T1w MRI images preprocessing steps included: *i)* intensity nonuniformity correction using the ANTS-based N4 algorithm;[4] *ii)* skull stripping using the FSL brain extraction algorithm (BET);[5] *iii)* tissue segmentation using FSL's FAST;[6] *iv)* spatial normalization using ANTS[7] to linearly and nonlinearly transform each native T1w image to a standardized MRI MNI152 space (2mm$^3$ isotropic). R-fMRI preprocessing comprised: *i)* realignment to anatomical images using AFNI *3dvolreg*[8] (functional co-registered to MNI 152 2mm$^3$) and *ii)* skull stripping using ANTS-based *3dAutomask* ; *iii)* estimation of Friston 24-motion parameters using AFNI and their linear and quadratic trends along with the mean framewise displacement (FD);[9, 10] *iv)* mean-based intensity normalization; *v)* nuisance signal regression. For the discovery analyses, nuisance signal regression included regressing out 24 motion parameters based on Friston 24-Parameter Model[9] and component-based noise correction (aCompCor).[11] aCompCor allows to regresse physiological noise indexed by the top five principal components within white matter and CSF masks.[11] As detailed in section 3.1, for the analyses on robustness to nuisance regression preprocessing, two different methods were used instead of aCompCor: global signal regression (GSR)[12] and 36 nuisance parameters regression as described below.[13] *vi)* Temporal bandpass filtering (i.e., removing frequencies outside the range of 0.01-0.1 Hz, using the AFNI *3dbandpass* algorithm) followed nuisance regression; then, *vii)* functional-to-anatomical coregistration using FSL FLIRT's boundary-based registration (BBR)[14] and spatial normalization of functional images to a the standard MNI152 space (2mm$^3$ isotropic) were obtained by applying linear and non-linear transforms from ANTs. *viii)* Lastly, spatial smoothing was applied using a 6mm full width at half maximum (FWHM) of a 3D Gaussian kernel using FSL.

##### *4.1 Robustness to distinct nuisance regression strategies*

To assess whether patterns of results from the discovery brain-behavior analyses were robust to different preprocessing pipelines, we repeated preprocessing using two validated nuisance regression pipelines[12, 13] as alternatives to aCompCor.[11] One pipeline included global signal regression (GSR)[12] along with 24-Friston model-based motion parameters, white matter and CSF regression. This was calculated at the individual level by averaging all voxel time-series within a whole-brain 25% probability FSL-based gray matter mask generated by CPAC and regressed out from each individual's time series. The other pipeline involved 36 parameter motion nuisance regression (36-P)[13] which regresses the primary 6 motion parameters, the white matter, CSF, and global time courses, their quadratic terms and temporal derivatives. Following preprocessing, robustness analyses were conducted at the cluster-level by 1) averaging the mean iFC values across all voxels in the masks of the cluster(s) showing significant iFC-behavior associations based on our discovery MDMR/SCA approach; and 2) correlating this mean iFC with the behavioral metric(s) of interest after regressing out age, sex assigned at birth, median FD, ADOS-2 CSS/ADHD-KSADS ratings consistent with our primary SCA analyses, as well as mean full brain connectivity of the given seed as in prior SCA studies.[15–17] we used the same approach to conduct the robustness assessment to longer time series or alternative behavioral measures in data processed with preprocessing pipeline used for primary analyses.

**5. KSADS-ADHD symptom severity**

Discovery brain-behavior analyses of ADHD symptoms were based on clinician's ratings on the *n*=18 KSADS-PL ADHD-specific questions that include *n*=9 questions on inattentive symptoms and *n*=9 on hyperactive/impulsive symptoms formulated on the basis of DSM-III_R and -IV ADHD symptom criteria.[18] The KSADS is a widely used and validated semi-structured diagnostic interview for children and adolescents in our study age range that assesses a wide range of psychopathology, including ADHD. Clinicians score each question using a 1-3 point rating scale, depending on the symptom presence, frequency and interference with functioning ("1"=absent, "2"=subthreshold, "3"=threshold). To attain a total score of ADHD symptom severity, we summed ratings across the 18 ADHD items after converting raw scores of "3" to 1, scores of "2" to 0.5, and scores of "1" to 0. We referred to this sum as the KSADS ADHD total score with a range between 0 and 18, with higher scores indicating greater severity. Scores of inattention and hyperactivity/impulsivity symptoms were derived by summing their 9 items separately (i.e., KSADS IN and KSADS H/I subscores; ranging from 0 to 9). To assess the validity of the KSADS ADHD clinician-based ratings, we correlated them with the SWAN-P averaged total, inattentive and hyperactivity/impulsivity ratings. All ratings were highly correlated, Pearson $r_{(165)}>0.5$ (ranging from $r=.54$ to $r=.63$), at $p<0.0001$, FDR corrected (see Figure S2).

**6. Gene expression decoding and enrichment analyses**

After *gene decoding- i.e., the* identification of genes whose expression in the brain is spatially correlated with the iFC map of interest, we run *gene enrichment*- i.e., testing if the expressed genes were enriched for genes previously reported to have greater rate of variants in ASD and ADHD.[19]

Consistent with prior work,[20, 21] we performed gene decoding analysis using NeuroVault,[22] a web-based repository that interoperates with the Allen Institute Human Brain (AHB) gene expression data.[23] The AHB is a multi-modal atlas that maps postmortem gene expression across the human brain integrating anatomic and genomic information from six donors; this has been widely used to establish inferential correlations between neuroimaging and transcriptomic brain maps.[23] Here, we focused on the iFC map of the MFG seed that showed a significant association to ADOS-2 CSS-Total based on our discovery MDMR/SCA framework. To assess the spatial similarity of each gene's spatial expression pattern with the MFG iFC, after uploading the MFG iFC map in NeuroVault (*NeuroVault ID*: 790618), we used mixed linear modeling as implemented in NeuroVault in each of the 6 donor brains across the 20,787 total genes included in the AHB atlas. The significance of the associations between the gene expression patterns and the MFG iFC map was then assessed via one-sample *t*-tests of the gene- and donor-specific slopes. The resulting list of genes from these t-tests were then thresholded for multiple comparisons (FDR at $q<0.05$). Given that positive and negative values were present among the voxels of the MFG iFC map, genes with positive and negative *t*-statistics were equally retained for gene enrichment analysis.

*Gene enrichment.* We conducted gene enrichment analysis using a list of *n*=1,046 unique genes selected from a large exome study of ASD and ADHD by Satterstrom et al.[19] The complete list of these 1,046 genes can be found in Table S4 and includes both protein-truncating and missense variants. Specifically, the list was composed of two sets of genes. One encompassed *n*=932 genes with high rate of protein truncating variants (PTV) and high probability (>0.9) of being loss-of-function intolerant (pLI), that were reported to have a similar rates of variance in both individuals without comorbid intellectual disability and diagnosis of autism and/or ADHD (so to be consistent with our sample characteristics).[19] The other set included genes representing PTVs with pLI<0.9, as well as Missense (Mis) variants, selected from the larger list of genes examined by Satterstrom et al.[19] and reported to also have a greater rate of variance in ADHD and ASD relative to neurotypical controls. Specifically, from the total of 1,514 genes not overlapping with the set of n=932 mentioned above, we selected *n*=114 based on the results of a two-tailed Fisher *z* exact test comparing the rate of variance between 'cases' (i.e., ASD and ADHD) without intellectual disability vs. controls (using the fisher.test function in R as in Satterstrom et al.).[19] These *n*=114 genes survived statistical significance at FDR corrected *q*<0.05. In order to reduce the risk of false findings with multiple gene enrichment tests, we did not assess other ASD- or ADHD-specific gene lists or recently reported shared common variants lists that only included < 30 genes.[24, 25] Enrichment was calculated by using hypergeometric statistical testing and quantified with odds ratios (OR). Consistent with prior work,[20, 21, 26, 27] the background total used for enrichment analyses was set to *n*=20,787 genes (i.e. the number of protein-coding genes quantified in the AHBn Atlas). Only those genes surviving correction for multiple comparisons at FDR *q*<0.05 were considered statistically significant. The custom R code used for genetic decoding and enrichment analysis is publicly available on GitHub at github.com/ChildMindInstitute/ASD_ADHD_BrainBehaviour_Genomics. Results from genetic enrichment analyses revealed significant enrichment for *n*=107 genes among the total *n*=1,519 genes topographically expressed within the MFG-iFC map. Of note, most of the enriched genes (*n*=100, 93%) represented protein-truncating variants, and were genes with high probability (>0.9) of being loss-of-function intolerant (*n*=101, 94%).

**7. Network Segregation Analyses**

Our primary discovery analyses revealed that internetwork iFC patterns between two nodes, one in the frontoparietal (FP) and one in the default mode (DMN) networks (i.e., MFG and PCC, respectively) were dimensionally related to autistic symptom severity. To explore the extent of internetwork iFC relationships with autistic symptoms beyond the two specific clusters identified in primary MDMR/SCA analyses (i.e., MFG and PCC), we computed the segregation index (SI)[28] across network pairs defined by the 7 functional Yeo network system.[29] As is Chan et al.,[30] SI was calculated to indicate the relative strength of *within*-network iFC compared to *between*-network iFC. Specifically, for the present study, *within*-network iFC was computed as the average node-to-node connectivity across all nodes within a functional network. *Between*-network iFC was computed as the average node-to-node connectivity between each node of a functional network and all nodes of the other functional networks. We examined all of Schaefer's 400 cortical parcellation atlas,[31] but 12 parcels located within limbic areas that were outside of our study specific brain mask (Figure S3). To calculate *within*- and *between*-network iFC across the remaining $n$=388 parcels, we extracted the mean time series from each parcel for each individual. We then interrogated the relationship between network pairs SI and autistic symptoms, by means of Pearson correlations between SI for each network pair and autistic symptoms indexed by ADOS-2 CSS-Total. Specifically, we quantified the SI of FP and DMN networks by assessing all parcels of these networks but excluding voxels that overlapped with MFG and PCC. Then, we also explored the SI of the FP with each of the remaining five networks pairs (i.e, FP-VIS, FP-MOT, FP-DAN, FP-VAN, FP-LIM), as well as the SI of the DMN with the same remaining five networks pairs (i.e., DMN-VIS, DMN-MOT, DMN-DAN, DMN-VAN, DMN-LIM). Of note, the SI across the parcels assigned to each pair of networks examined was computed for each individual averaging across hemispheres. Results were corrected for multiple comparisons at FDR $q$<0.05 (see Figure 2c).

**8. Control genetic decoding/enrichment analysis**

To examine the specificity of the genetic enrichment results, we performed a control analysis. Specifically, we decoded genes expressed in an iFC map derived by a control seed and tested their enrichment with the same ASD/ADHD shared list used in our main enrichment analyses. To carry out this approach, we first selected a control seed based on the following criteria: i) falling in an area with low and non-significant brain-behavior statistics (i.e., Z>-0.5 and Z<0.5) in the MDMR analysis of ADOS-2 CSS, ii) seed with an iFC map with minimal spatial correlations with the MFG iFC map (Pearson $r$<=-0.20). The resulting seed was placed in the supracalcarine cortex (x=22, y=12, z=22; 5mm radius sphere; NeuroVault: ID 781986) of the central visual functional network. Then, we used the derived iFC map from this control seed to carry out gene decoding and gene enrichment analysis as described in main text and above in section 6. Analyses revealed that the $n$=7,572 brain decoded genes from this central visual iFC map (full list available in GitHub) were not significantly enriched for the ASD/ADHD genes tested in main analysis (OR=1.7, $p$=0.2$^{ns}$). Overall, these results suggest that the ASD/ADHD gene enrichment findings are specific to the MFG-iFC map relevant for autism symptoms across diagnoses.

**9. Gene ontology analysis**

To explore the biological processes associated with the *n*=107 genes resulting from the genetic enrichment analyses, we used the Gene Ontology database implemented with AmiGO 2. AmiGO 2 is a web-based open-source set of tools for querying, browsing, and visualizing GO data that includes electronic and manually curated annotations of GO biological processes.[32] We used the 2024-01-17 release version of the GO database.[33] Specifically, we compared our list of 107 genes to a reference list of all genes in the database for *Homo Sapiens* using over-representation analysis. All GO terms surviving FDR correction at $q<0.05$ were retained and they are listed in Table S7, including the full name of the related GO biological process terms. Given that biological processes are highly related and to aid interpretation of results, GO sorts terms hierarchically starting from the most specific subclass to the more general terms (referred to as parent terms). These general terms are shown as indented terms directly below the more specific terms in Table S7. Given that the most specific terms (or "child" terms) out of the top-20 most statistically significant terms indexed processes related to neuron morphogenesis, Figure S6 summarizes the most specific terms in this subclass, along with their GO term identification, number and definitions. These include axonogenesis, dendritic spine morphogenesis, axon extension, and axon guidance, all of which share neuron projection morphogenesis as a 'parent' term (also see Figure 4).

### SUPPLEMENTARY FIGURES

**Supplementary Figure S1: Diagram of the preprocessing steps for T1-weighted and resting-state fMRI data. a)** Diagram showing the preprocessing steps for T1-weighted (T1w) structural MRI scans (left) and for resting-state fMRI scans (right) used for all analyses; **b)** the inset illustrates specific details on the three different nuisance regression strategies used for discovery and robustness analysis (i.e., a principal components analysis strategy (aCompCor), global signal regression (GSR), or a 36-motion parameter model strategy (36P)).

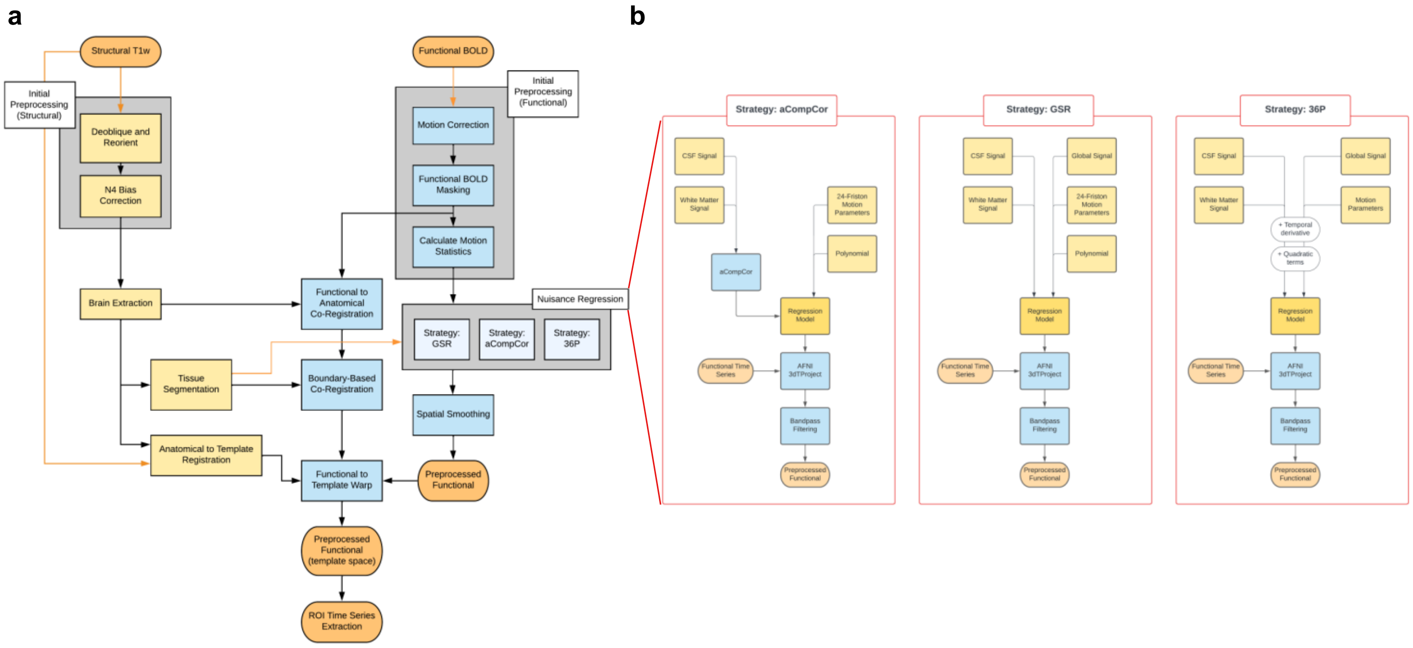

**Supplementary Figure S2: Correlations between KSADS and SWAN ADHD symptoms.** The scatterplots show the correlation between clinician-based KSADS total, inattentive and hyperactive/impulsive subscore distributions, and the corresponding parent-based SWAN (from left to right) across the *n*=166 children included in this study. Abbreviations: KSADS, Kiddie-Schedule for Affective Disorders and Schizophrenia for School-Age Children–Present and Lifetime Version; SWAN, Strengths and Weaknesses of ADHD Symptoms and Normal Behavior.

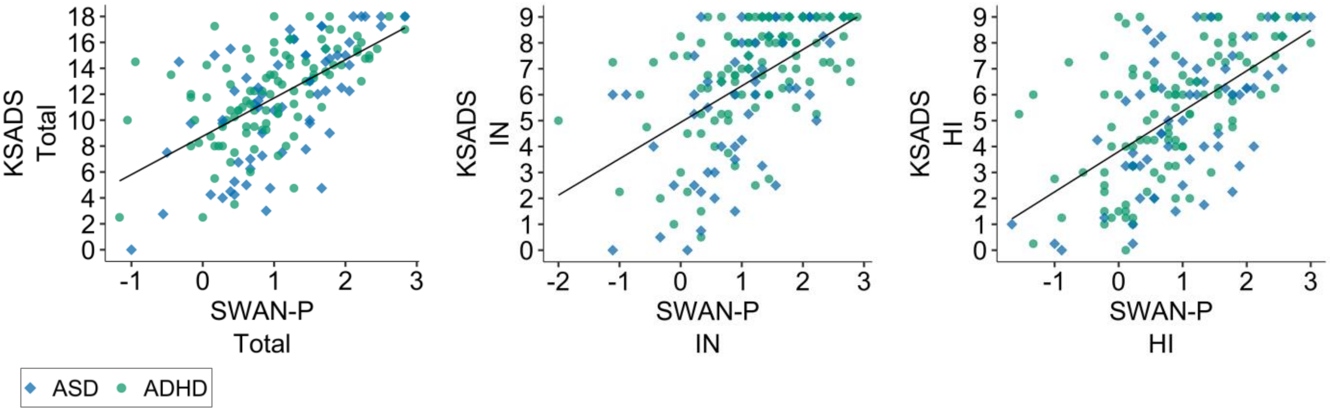

**Supplementary Figure S3**: **Group-level study-specific mask.** MNI 152 standard brain overlaid with a 100% group-level whole-brain functional volume mask (shown in yellow), i.e., including voxels present across all *n*=166 participants (total number of voxels, *n*=10,664).

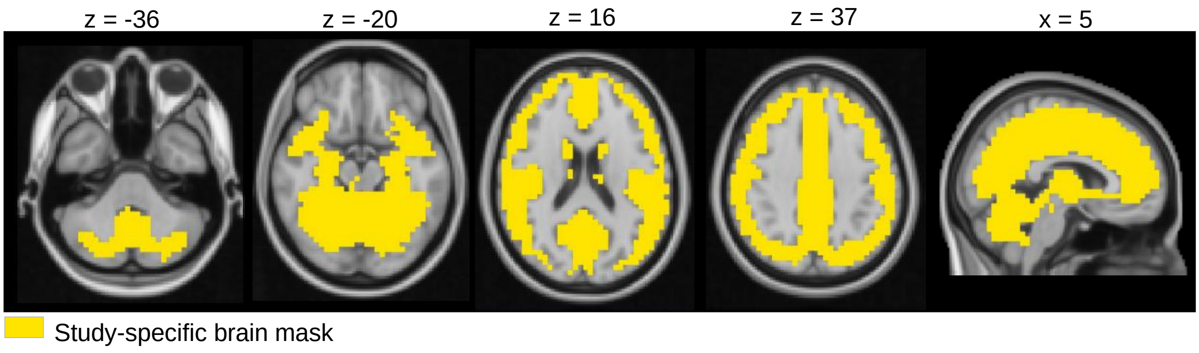

**Supplementary Figure S4: ADOS-2 symptom subdomains association with iFC. a)** Z-score voxel-level maps resulting from Multivariate Distance Matrix Regression (MDMR) using calibrated severity scores for social affect (CSS-SA) or restricted repetitive behaviors (CSS-RRB) in the model. At the whole-brain level, no associations survived Gaussian Random Field Theory correction ($Z>3.1$, $p\leq0.01$). However, consistent with notable subthreshold iFC associations with SA in PCC, correlations conducted at the cluster-level based on the findings obtained in the primary analysis using CSS-Total, revealed a moderate association with CSS-SA ($r_{(164)}$=0.23, $p$=0.003) but not with CSS-RRB ($r_{(164)}$=0.08, $p$=0.29). This is shown in the **b)** scatter plots of relationships between CSS-SA (left) or CSS-RRB (right) and MFG iFC with PCC across children with autism spectrum disorder (ASD; blue diamonds) and ADHD without ASD ($ADHD_{w/oASD}$; green circles). Both iFC and ADOS-2 CSS-SA and CSS-RRB are shown as residuals after regressing the same covariates used in discovery brain-behavior analyses (i.e., median FD, age, sex and KSADS ADHD total score). Z-score maps and the circuit examined were overlaid onto brain surface maps.

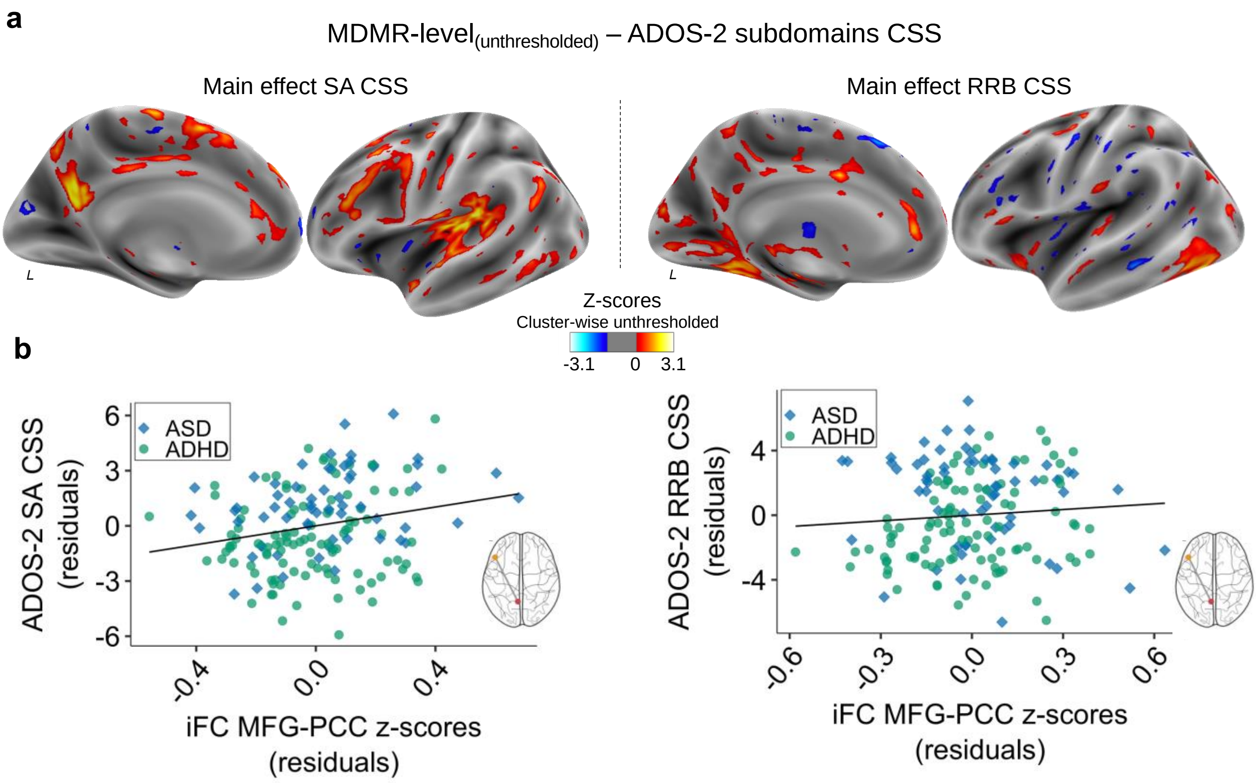

**Supplementary Figure S5: MDMR analysis revealed no significant brain-behavior relationships for ADHD symptoms transdiagnostically.** The Z-score maps show, at the voxel level, the correlations between iFC and ADHD total, inattentive, and hyperactive impulsive symptoms (going from left to right) indexed by **a)** clinician-based KSADS ratings and **b)** parent-based SWAN ratings. There were no statistically significant correlations after cluster-level Gaussian Field Theory correction at Z>3.1, *p*≤0.01.

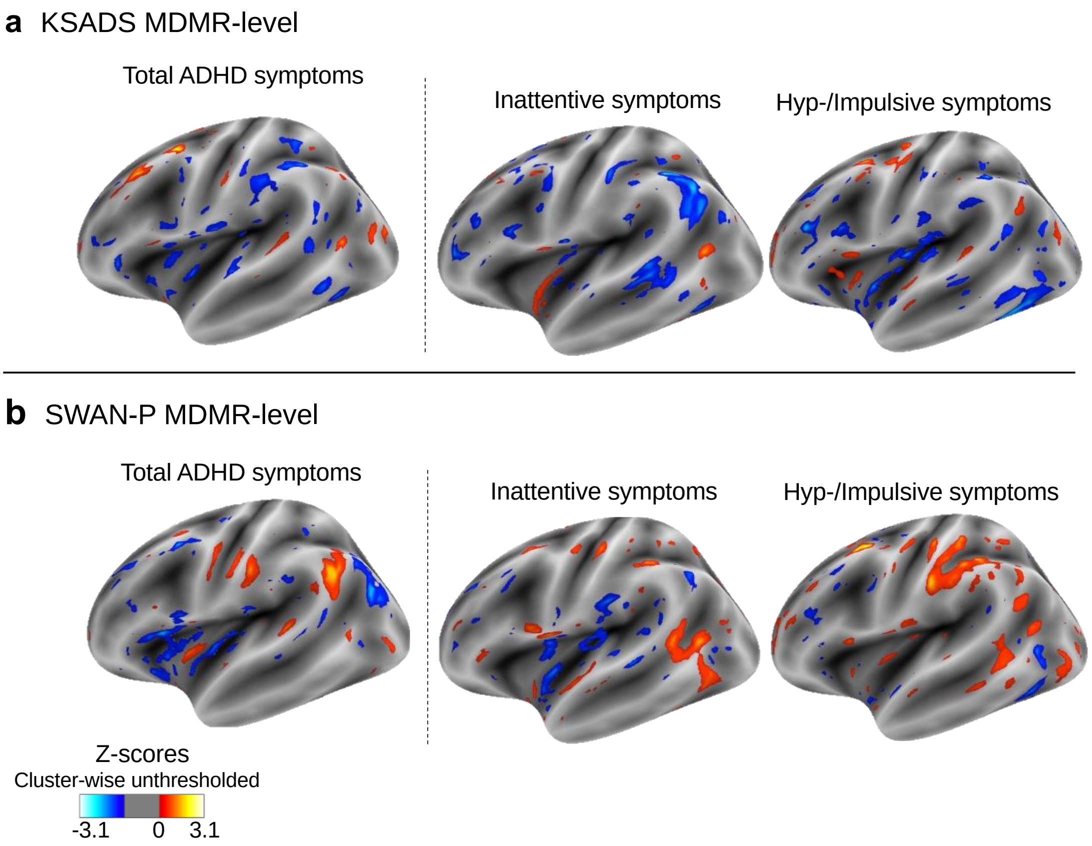

**Supplementary Figure S6: Hierarchy tree of the most representative Gene Ontology (GO) biological processes terms for the top statistically significant results.** Exploratory analysis leveraging the GO annotation resource[32] allowed us to identify associations between biological process terms and the *n*=107 genes identified as enriched in the iFC map relevant for autistic traits. GO biological process terms surviving FDR correction at $q<0.05$ were retained (see Table S7 and Supplementary material). GO biological processes are organized in a hierarchy from top to bottom of a hierarchy tree, going from more general to increasingly more specific processes (i.e., "parent" and 'child" respectively). Here, we visualize the hierarchy tree associated with the most specific of the top-most statistically significant GO terms. All of them were related to the 'parent' term "*Neuron projection morphogenesis*", shown in the dark gray layer at the top of the tree. The lower layers of the figure in lighter gray show the related 'child' terms (i.e., *"Axonogenesis", "Dendritic spine morphogenesis", "Axon extension", "Axon guidance"* and *"Regulation of axonogenesis"*). For each GO term, the term name and the number of genes showing significant associations with the GO term are indicated, as well as the FDR corrected p-values at q<0.05. The GO term IDs are also shown in blue boxes. A definition of each GO term provided by the GO database,[32] is as follows. **a. Neuron projection morphogenesis:** the process in which the anatomical structures of a neuron projection are generated and organized. A neuron projection is any process extending from a neural cell, such as axons or dendrites. **b. Axonogenesis:** *de novo* generation of a long process of a neuron, including the terminal branched region. Refers to the morphogenesis or creation of shape or form of the developing axon, which carries efferent (outgoing) action potentials from the cell body towards target cells. **c. Dendritic spine morphogenesis:** The process in which the anatomical structures of a dendritic spine are generated and organized. A dendritic spine is a protrusion from a dendrite and a specialized subcellular compartment involved in synaptic transmission. **d. Axon extension:** Long distance growth of a single axon process involved in cellular development. **e. Axon guidance:** the chemotaxis process that directs the migration of an axon growth cone to a specific target site in response to a combination of attractive and repulsive cues. **f. Regulation of axonogenesis:** any process that modulates the frequency, rate or extent of axonogenesis, the generation of an axon, the long process of a neuron.

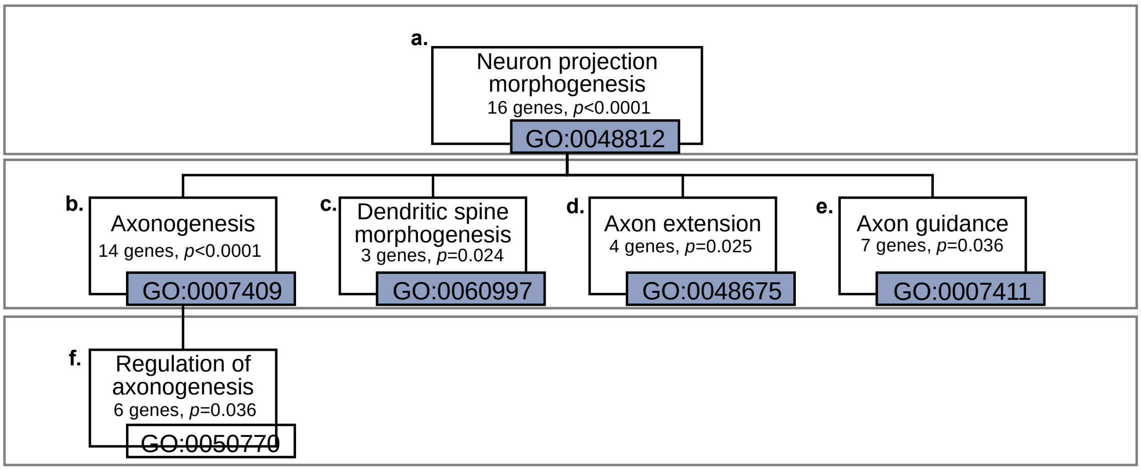

**Table S1.** Sample characteristics by phenotyping site (NYU vs CMI).

| Variable | NYU | CMI | Statistics | | | |
|---|---|---|---|---|---|---|
| | n=81 | n=85 | df | W $\chi 2$ | p value | FDR-p value |
| **Primary diagnosis,** # (%) | | | 1 | 0.01 | 0.94 | 0.94 |
| ASD | 30 (37) | 33 (39) | | | | |
| $ADHD_{w/o\ ASD}$ | 51 (63) | 52 (61) | | | | |
| **Age**, years, M (SD) | 8.8 (1.8) | 8.9 (1.7) | - | 3575.00 | 0.67 | 0.86 |
| **Sex assigned at birth,** males #, (%) | 62 (77) | 63 (74) | 1 | 0.03 | 0.86 | 0.97 |
| **Ethnicity,** Hispanic #, (%)[a] | 20 (25) | 19 (23) | 1 | 0.02 | 0.90 | 0.97 |
| **Race,** #, (%)[a,b] | | | 4 | 6.40 | 0.17 | 0.58 |
| White | 51 (63) | 44 (52) | | | | |
| Mixed | 15 (19) | 11 (13) | | | | |
| Black/African American | 8 (10) | 11 (13) | | | | |
| Asian | 4 (5) | 10 (12) | | | | |
| Other | 3 (4) | 8 (10) | | | | |
| **Socio-economic status,** class #, (%)[c] | | | 1 | 0.00 | 0.99 | 0.99 |
| Classes 1-3 | 21 (27) | 24 (29) | | | | |
| Classes 4-5 | 56 (73) | 60 (71) | | | | |
| **Psychoactive Medication Status,** #, (%)[d] | | | 2 | 1.59 | 0.45 | 0.80 |
| Med naïve | 56 (71) | 61 (72) | | | | |
| Currently on medication | 19 (24) | 16 (19) | | | | |
| Not Naïve but current off treatment | 4 (5) | 8 (9) | | | | |
| **Psychiatric Comorbidity** (yes) **#, (%)** | 46 (57) | 62 (73) | 1 | 4.08 | 0.04 | 0.31 |
| **DAS-II IQ Standard scores**, M (SD) | | | | | | |
| Verbal IQ | 105.3 (15.7) | 108.5 (16.6) | - | 3834.50 | 0.21 | 0.58 |
| Non-verbal IQ | 100.9 (18.9) | 106.2 (16.8) | - | 4045.00 | 0.05 | 0.31 |
| Full-scale IQ | 102.7 (16.1) | 106.2 (17.1) | - | 3819.50 | 0.22 | 0.58 |
| **ADOS-2**, M (SD) | | | | | | |
| Expressive Language[a] | 7.9 (0.3) | 7.9 (0.5) | - | 3604.50 | 0.33 | 0.75 |
| Social affect CSS | 5.4 (2.4) | 4.9 (2.7) | - | 2993.50 | 0.14 | 0.58 |
| RRB CSS | 4 (2.9) | 5.2 (3.4) | - | 4196.50 | 0.01 | 0.26 |
| Total CSS | 4.8 (2.6) | 4.6 (2.8) | - | 3260.00 | 0.55 | 0.80 |
| **K-SADS ADHD severity**, M (SD) | | | | | | |
| Inattentive | 6.7 (2.4) | 6.9 (2) | - | 3408.00 | 0.91 | 0.97 |
| Hyperactive | 5.5 (2.6) | 5.8 (2.3) | - | 3244.50 | 0.52 | 0.80 |
| Total | 12.2 (4) | 12.7 (3.5) | - | 3253.50 | 0.54 | 0.80 |
| **In-scanner motion**, median FD, M (SD) | 0.1 (0) | 0.1 (0) | - | 3697.50 | 0.41 | 0.80 |

Over the course of the study, the enrolling and phenotyping site transferred from the NYU Grossman School of Medicine Child Study Center to the Child Mind Institute (CMI) where the principal investigator (ADM) acquired a new position. Wilcoxon Rank sum tests were used to calculate between site differences for the continous variables and Pearson Chi-square tests for categorical variables. All the obtained p-values were corrected using False Disocvery Rate (FDR) at q>0.05. **a.** Missing for one child enrolled at the site; **b.** "Other" includes American Indian/Alaskan Native, Native Hawaiian/Pacific Islander, mixed race or other not specified. Of the n=3 children from NYU under "Other", 1 identified as American Indian/Alaskan Native, and 2 as 'other.' Of the 13 children with ADHD under other, 1 identified as Native Hawaiian/other Pacific Islander, 7 as 'other'; **c.** The group enrolled at thge New York University (NYU) site included: n=4 children in the socioeconomic class (SES) Class 1, n=6 in SES Class 2, n=11 in SES Class 3, n=16 in SES Class 4, and n=40 in SES Class 5. The CMI group included n=5 children in SES Class 1, n=6 children in SES Class 2, n=13 in SES Class 3, n=21 in SES Class 4 and n=39 SES Class 5. Missing data for n=4 children from NYU and n=1 from CMI sites; **d.** Information on medication status was not available for n=2 children from the NYU site; **e.** Autism Diagnostic Observation Schedule second edition(ADOS-2) based measure of expressive language validated in Mazurek et al. 2019, it ranges from 1 to 8 ("1" corresponds to "No spontaneous words or word approximations" and "8" to "Uses sentences in a largely correct fashion (must use some complex speech)". **Abbreviations:** ASD, Autism Spectrum Disorder; ADHDw/oASD, Attention Deficit Hyperactivity Disorder withouht ASD; DAS-II, Differential Ability Scale-2nd Edition; CSS, calibrated severity scores; K-SADS; Kiddie Schedule for Affective Disorders and Schizophrenia; df, degrees of freedom; W $\chi 2$, Wilcoxon /chi-square statistic.

**Table S2.** MRI imaging parameters per protocol.

| Scan | Duration | Matrix | SI | FOV | FOV Phase | Res | TR | TE | TI | FA | MBA | Vol |
|---|---|---|---|---|---|---|---|---|---|---|---|---|
| | min:sec | | # | | % | mm | ms | ms | ms | degrees | | |
| **T1w** | 6:36 | 320x300 | 208 | 256x256 | 93.8 | 0.8x0.8x0.8 | 2400 | 2.24 | 1060 | 8 | - | - |
| **R-fMRI - 1** | 6:20 | 90x92 | 66 | 216x221 | 102.2 | 2.4x2.4x2.4 | 800 | 30 | - | 55 | 6 | 465 |
| **R-fMRI - 2** | 4:39 | 90x92 | 66 | 216x221 | 102.2 | 2.4x2.4x2.4 | 800 | 30 | - | 55 | 6 | 339 |

MRI imaging parameters at each scan in the protocol. Abbreviations: FA, flip angle; FOV, field of view; MBA, multiband acceleration; TE, echo time; TI, inversion time; TR, repetition time; T1-w, T1-weighted (MPRAGE); R-fMRI, resting state functional MRI; Res, spatial resolution; SI, slice #; Vol, number of volumes.

**Table S3.** Sample characteristics of children included vs. those excluded from the present analyses.

| Variable | Included | Excluded after MRI QA | Statisitics | | | |
|---|---|---|---|---|---|---|
| | n=166 | n=28 | df | W $\chi 2$ | p value | FDR-p value |
| **Primary diagnosis,** # (%) | | | 1 | 1.10 | 0.29 | 0.63 |
| ASD | 63 (38) | 14 (50) | | | | |
| $ADHD_{w/o\ ASD}$ | 103 (62) | 14 (50) | | | | |
| **Age**, years, M (SD) | 8.9 (1.7) | 8.7 (2.0) | - | 2380.5 | 0.84 | 0.95 |
| **Sex assigned at birth,** males #, (%) | 125 (75) | 22 (81) | 2 | 0.61 | 0.74 | 0.95 |
| **Ethnicity,** Hispanic #, (%)[a] | 39 (24) | 10 (37) | 1 | 1.54 | 0.21 | 0.63 |
| **Race,** #, (%)[a,b] | | | 4 | 4.60 | 0.33 | 0.63 |
| White | 95 (58) | 14 (52) | | | | |
| Mixed | 26 (16) | 5 (19) | | | | |
| Black/African American | 19 (12) | 6 (22) | | | | |
| Asian | 14 (8) | 0 (0) | | | | |
| Other | 11 (7) | 2 (7) | | | | |
| **Socio-economic status,** class #, (%)[c] | | | 1 | 0.00 | 1.00 | 1.00 |
| Classes 1-3 | 45 (28) | 6 (27) | | | | |
| Classes 4-5 | 116 (72) | 16 (73) | | | | |
| **DAS-II IQ Standard scores**, M (SD) | | | | | | |
| Verbal IQ | 107 (16) | 102 (13) | - | 2800.50 | 0.04 | 0.63 |
| Non-verbal IQ | 104 (18) | 102 (17) | - | 2298.00 | 0.83 | 0.95 |
| Full-scale IQ | 105 (17) | 101 (14) | - | 2439.50 | 0.46 | 0.79 |
| **ADOS-2**, M (SD) | | | | | | |
| Expressive Language[d] | 7.9 (0.4) | 7.8 (0.4) | - | 2558.00 | 0.14 | 0.63 |
| Social affect CSS | 5.1 (2.6) | 5.8 (2.6) | - | 1874.50 | 0.14 | 0.63 |
| RRB CSS | 4.6 (3.2) | 5.2 (3.0) | - | 2015.00 | 0.33 | 0.63 |
| Total CSS | 4.7 (2.7) | 5.4 (2.6) | - | 1884.50 | 0.15 | 0.63 |
| **K-SADS ADHD severity**, M (SD) | | | | | | |
| Inattentive | 6.8 (2.2) | 7.1 (1.8) | - | 2009.00 | 0.97 | 1.00 |
| Hyperactive | 5.6 (2.4) | 5.5 (2.8) | - | 1943.00 | 0.78 | 0.95 |
| Total | 12.4 (3.8) | 11.3 (5.1) | - | 2479 | 0.574 | 0.886 |

Wilcoxon Rank sum tests were used to calculate between group differences for the continous variables and Pearson Chi-square tests for categorical variables. p-values were corrected using False Disocvery Rate (FDR) at q>0.05, p values significant after correction are shown in bold in the FDR p-value column. **a.** Missing from n=1 child from the included group, and n=1 from the excluded group; **b.** "Other" includes American Indian/Alaskan Native, Native Hawaiian/Pacific Islander, mixed race or other not specified. Of the n=11 children from the included group under "Other", 1 identified as American Indian/Alaskan Native,1 identified as Native Hawaiian/other Pacific Islander, and 9 as 'other'. Of the n=2 children from the excluded group under "Other",identified as 'other'; **c.** The included group included: n=9 children in socioeconomic class (SES) Class 1, n=12 in SES Class 2, n=24 in SES Class 3, n=37 in SES Class 4, and n=79 in SES Class 5. The excluded group included n=3 children in SES Class 2, n=3 in SES Class 3, n=7 in SES Class 4 and n=9 SES Class 5. Missing data for n=5 children from the included group and n=6 from the excluded one; **d.** Autism Diagnostic Observation Schedule second edition (ADOS-2) based measure of expressive language validated in Mazurek et al. 2019, it ranges from 1 to 8 ("1" corresponds to "No spontaneous words or word approximations" and "8" to "Uses sentences in a largely correct fashion (must use some complex speech)". **Abbreviations:** ASD, Autism Spectrum Disorder; ADHDw/oASD, Attention Deficit Hyperactivity Disorder wihtout ASD; DAS-II, Differential Ability Scales-2nd Edition; CSS, calibrated severity scores; K-SADS; Kiddie Schedule for Affective Disorders and Schizophrenia; df, degrees of freedom; W $\chi 2$, Wilcoxon /chi-square statistic

**Table S4.** Curated list of *n*=1,046 genes tested for genetic enrichment in the present study.

| Gene symbol | Gene name |
|---|---|
| AAK1 | AP2 associated kinase 1 |
| ABCD3 | ATP-binding cassette, sub-family D (ALD), member 3 |
| ABCF2 | ATP-binding cassette, sub-family F (GCN20), member 2 |
| ABHD16A | Abhydrolase Domain Containing 16A, Phospholipase |
| ABLIM1 | actin binding LIM protein 1 |
| ABR | active BCR-related |
| ACACA | acetyl-CoA carboxylase alpha |
| ACLY | ATP citrate lyase |
| ACTN1 | actinin, alpha 1 |
| ACTN2 | actinin, alpha 2 |
| ADAD1 | adenosine deaminase domain containing 1 (testis-specific) |
| ADAM10 | ADAM metallopeptidase domain 10 |
| ADAM17 | ADAM metallopeptidase domain 17 |
| ADCY1 | adenylate cyclase 1 (brain) |
| ADCY9 | adenylate cyclase 9 |
| AFF3 | AF4/FMR2 family, member 3 |
| AGAP1 | ArfGAP with GTPase domain, ankyrin repeat and PH domain 1 |
| AGO2 | Argonaute RISC Catalytic Component 2 |
| AGO3 | Argonaute RISC Catalytic Component 3 |
| AGO4 | Argonaute RISC Catalytic Component 4 |
| AHCYL2 | adenosylhomocysteinase-like 2 |
| AHR | aryl hydrocarbon receptor |
| AKT2 | v-akt murine thymoma viral oncogene homolog 2 |
| ALDH1A1 | aldehyde dehydrogenase 1 family, member A1 |
| ANAPC7 | anaphase promoting complex subunit 7 |
| ANK1 | ankyrin 1, erythrocytic |
| ANK2 | ankyrin 2, neuronal |
| ANK3 | ankyrin 3, node of Ranvier (ankyrin G) |
| ANKFY1 | ankyrin repeat and FYVE domain containing 1 |
| ANKRD11 | ankyrin repeat domain 11 |

| | |
|---|---|
| ANKRD12 | ankyrin repeat domain 12 |
| ANKRD34A | ankyrin repeat domain 34A |
| ANKS1B | ankyrin repeat and sterile alpha motif domain containing 1B |
| ANO8 | anoctamin 8 |
| AP2B1 | adaptor-related protein complex 2, beta 1 subunit |
| AP2M1 | adaptor-related protein complex 2, mu 1 subunit |
| AP2S1 | adaptor-related protein complex 2, sigma 1 subunit |
| AP3B1 | adaptor-related protein complex 3, beta 1 subunit |
| AP3B2 | adaptor-related protein complex 3, beta 2 subunit |
| AP4E1 | adaptor-related protein complex 4, epsilon 1 subunit |
| APBB1 | amyloid beta (A4) precursor protein-binding, family B, member 1 (Fe65) |
| APPBP2 | amyloid beta precursor protein (cytoplasmic tail) binding protein 2 |
| AQR | aquarius homolog (mouse) |
| ARAP3 | ArfGAP with RhoGAP domain, ankyrin repeat and PH domain 3 |
| ARFGEF1 | ADP-ribosylation factor guanine nucleotide-exchange factor 1 (brefeldin A-inhibited) |
| ARFGEF2 | ADP-ribosylation factor guanine nucleotide-exchange factor 2 (brefeldin A-inhibited) |
| ARFIP2 | ADP-ribosylation factor interacting protein 2 |
| ARHGAP26 | Rho GTPase activating protein 26 |
| ARHGAP32 | Rho GTPase Activating Protein 32 |
| ARHGAP5 | Rho GTPase activating protein 5 |
| ARHGEF1 | Rho guanine nucleotide exchange factor (GEF) 1 |
| ARHGEF17 | Rho guanine nucleotide exchange factor (GEF) 17 |
| ARHGEF2 | Rho/Rac guanine nucleotide exchange factor (GEF) 2 |
| ARHGEF3 | Rho guanine nucleotide exchange factor (GEF) 3 |
| ARID1B | AT rich interactive domain 1B (SWI1-like) |
| ARID4A | AT rich interactive domain 4A (RBP1-like) |
| ARID4B | AT rich interactive domain 4B (RBP1-like) |
| ARID5B | AT rich interactive domain 5B (MRF1-like) |
| ARIH1 | ariadne homolog, ubiquitin-conjugating enzyme E2 binding protein, 1 (Drosophila) |
| ASAP1 | ArfGAP with SH3 domain, ankyrin repeat and PH domain 1 |
| ASH1L | ash1 (absent, small, or homeotic)-like (Drosophila) |

| | |
|---|---|
| ASH2L | ash2 (absent, small, or homeotic)-like (Drosophila) |
| ASTN1 | astrotactin 1 |
| ASUN | asunder spermatogenesis regulator |
| ASXL3 | additional sex combs like 3 (Drosophila) |
| ATAD2 | ATPase family, AAA domain containing 2 |
| ATAD2B | ATPase family, AAA domain containing 2B |
| ATG13 | autophagy related 13 |
| ATG2A | autophagy related 2A |
| ATG4B | autophagy related 4B, cysteine peptidase |
| ATP11A | ATPase, class VI, type 11A |
| ATP13A1 | ATPase type 13A1 |
| ATP13A3 | ATPase type 13A3 |
| ATP1A1 | ATPase, Na+/K+ transporting, alpha 1 polypeptide |
| ATP1A3 | ATPase, Na+/K+ transporting, alpha 3 polypeptide |
| ATP2B1 | ATPase, Ca++ transporting, plasma membrane 1 |
| ATP2B2 | ATPase, Ca++ transporting, plasma membrane 2 |
| ATP8B2 | ATPase, aminophospholipid transporter, class I, type 8B, member 2 |
| ATRN | attractin |
| ATXN2 | ataxin 2 |
| ATXN2L | ataxin 2-like |
| B4GALT5 | UDP-Gal:betaGlcNAc beta 1,4- galactosyltransferase, polypeptide 5 |
| BACE1 | beta-site APP-cleaving enzyme 1 |
| BAG6 | BAG cochaperone 6 |
| BAI2 | brain-specific angiogenesis inhibitor 2 |
| BAI3 | brain-specific angiogenesis inhibitor 3 |
| BAZ1B | bromodomain adjacent to zinc finger domain, 1B |
| BAZ2A | bromodomain adjacent to zinc finger domain, 2A |
| BAZ2B | bromodomain adjacent to zinc finger domain, 2B |
| BBX | bobby sox homolog (Drosophila) |
| BCAR3 | breast cancer anti-estrogen resistance 3 |
| BCL9L | B-cell CLL/lymphoma 9-like |
| BCR | breakpoint cluster region |
| BMS1 | BMS1 homolog, ribosome assembly protein (yeast) |
| BNC2 | basonuclin 2 |

| | |
|---|---|
| BOD1L1 | Biorientation Of Chromosomes In Cell Division 1 Like 1 |
| BPTF | bromodomain PHD finger transcription factor |
| BRD2 | bromodomain containing 2 |
| BRD7 | bromodomain containing 7 |
| BRINP1 | BMP/Retinoic Acid Inducible Neural Specific 1 |
| BRINP2 | BMP/Retinoic Acid Inducible Neural Specific 2 |
| BRPF3 | bromodomain and PHD finger containing, 3 |
| BRWD1 | bromodomain and WD repeat domain containing 1 |
| BRWD3 | bromodomain and WD repeat domain containing 3 |
| BSN | bassoon (presynaptic cytomatrix protein) |
| BTBD11 | BTB (POZ) domain containing 11 |
| C10orf12 | chromosome 10 open reading frame 12 |
| C10orf2 | chromosome 10 open reading frame 2 |
| C11orf57 | chromosome 11 open reading frame 57 |
| C20orf112 | chromosome 20 open reading frame 112 |
| C7orf55-LUC7L2 | chromosome 7 open reading frame 55 and LUC7-like 2 |
| CA10 | carbonic anhydrase X |
| CABP1 | calcium binding protein 1 |
| CACNA1B | calcium channel, voltage-dependent, N type, alpha 1B subunit |
| CACNA1D | calcium channel, voltage-dependent, L type, alpha 1D subunit |
| CACNA1E | calcium channel, voltage-dependent, R type, alpha 1E subunit |
| CACNA1G | calcium channel, voltage-dependent, T type, alpha 1G subunit |
| CACNA2D2 | calcium channel, voltage-dependent, alpha 2/delta subunit 2 |
| CACNA2D3 | calcium channel, voltage-dependent, alpha 2/delta subunit 3 |
| CACNB1 | calcium channel, voltage-dependent, beta 1 subunit |
| CACNG3 | calcium channel, voltage-dependent, gamma subunit 3 |
| CACTIN | Cactin, Spliceosome C Complex Subunit |
| CAD | carbamoyl-phosphate synthetase 2, aspartate transcarbamylase, and dihydroorotase |
| CADM1 | cell adhesion molecule 1 |
| CADM2 | cell adhesion molecule 2 |

| | |
|---|---|
| CADPS2 | Ca++-dependent secretion activator 2 |
| CALCRL | calcitonin receptor-like |
| CAMSAP2 | Calmodulin Regulated Spectrin Associated Protein Family Member 2 |
| CAMSAP3 | Calmodulin Regulated Spectrin Associated Protein Family Member 3 |
| CAMTA2 | calmodulin binding transcription activator 2 |
| CBX8 | chromobox homolog 8 |
| CCDC88A | coiled-coil domain containing 88A |
| CCNJ | cyclin J |
| CCT5 | chaperonin containing TCP1, subunit 5 (epsilon) |
| CCT6A | chaperonin containing TCP1, subunit 6A (zeta 1) |
| CCZ1 | CCZ1 Homolog, Vacuolar Protein Trafficking And Biogenesis Associated |
| CDAN1 | codanin 1 |
| CDC42BPA | CDC42 binding protein kinase alpha (DMPK-like) |
| CDH9 | cadherin 9, type 2 (T1-cadherin) |
| CDYL | chromodomain protein, Y-like |
| CELF4 | CUGBP Elav-Like Family Member 4 |
| CELSR1 | cadherin, EGF LAG seven-pass G-type receptor 1 (flamingo homolog, Drosophila) |
| CELSR2 | cadherin, EGF LAG seven-pass G-type receptor 2 (flamingo homolog, Drosophila) |
| CERS2 | Ceramide Synthase 2 |
| CFH | complement factor H |
| CHD3 | chromodomain helicase DNA binding protein 3 |
| CHD5 | chromodomain helicase DNA binding protein 5 |
| CHD6 | chromodomain helicase DNA binding protein 6 |
| CHD8 | chromodomain helicase DNA binding protein 8 |
| CHD9 | chromodomain helicase DNA binding protein 9 |
| CHMP7 | charged multivesicular body protein 7 |
| CHRM3 | cholinergic receptor, muscarinic 3 |
| CIC | capicua homolog (Drosophila) |
| CIT | citron (rho-interacting, serine/threonine kinase 21) |
| CIZ1 | CDKN1A interacting zinc finger protein 1 |
| CKAP5 | cytoskeleton associated protein 5 |
| CLASP2 | cytoplasmic linker associated protein 2 |
| CLCN7 | chloride channel, voltage-sensitive 7 |

| | |
|---|---|
| CLIP2 | CAP-GLY domain containing linker protein 2 |
| CLPTM1 | cleft lip and palate associated transmembrane protein 1 |
| CLSPN | claspin |
| CMTR1 | Cap Methyltransferase 1 |
| CNOT10 | CCR4-NOT transcription complex, subunit 10 |
| CNOT3 | CCR4-NOT transcription complex, subunit 3 |
| CNTN1 | contactin 1 |
| CNTN4 | contactin 4 |
| COBL | cordon-bleu homolog (mouse) |
| COL11A2 | collagen, type XI, alpha 2 |
| COL12A1 | collagen, type XII, alpha 1 |
| COL27A1 | collagen, type XXVII, alpha 1 |
| COL2A1 | collagen, type II, alpha 1 |
| COL4A3BP | collagen, type IV, alpha 3 (Goodpasture antigen) binding protein |
| COLEC12 | collectin sub-family member 12 |
| COPG1 | COPI Coat Complex Subunit Gamma 1 |
| COPS5 | COP9 constitutive photomorphogenic homolog subunit 5 (Arabidopsis) |
| COPS7A | COP9 constitutive photomorphogenic homolog subunit 7A (Arabidopsis) |
| CPE | carboxypeptidase E |
| CPNE6 | copine VI (neuronal) |
| CPT1A | carnitine palmitoyltransferase 1A (liver) |
| CREB1 | cAMP responsive element binding protein 1 |
| CRTC2 | CREB regulated transcription coactivator 2 |
| CSF1R | colony stimulating factor 1 receptor |
| CSMD3 | CUB and Sushi multiple domains 3 |
| CSNK1D | casein kinase 1, delta |
| CTIF | Cap Binding Complex Dependent Translation Initiation Factor |
| CTNNA2 | catenin (cadherin-associated protein), alpha 2 |
| CTNND1 | catenin (cadherin-associated protein), delta 1 |
| CTNND2 | catenin (cadherin-associated protein), delta 2 (neural plakophilin-related arm-repeat protein) |
| CTPS1 | CTP Synthase 1 |
| CTR9 | Ctr9, Paf1/RNA polymerase II complex component, homolog (S. cerevisiae) |

| | |
|---|---|
| CUL2 | cullin 2 |
| CUL5 | cullin 5 |
| CUL9 | cullin 9 |
| CUX2 | cut-like homeobox 2 |
| CYFIP1 | cytoplasmic FMR1 interacting protein 1 |
| DAAM1 | dishevelled associated activator of morphogenesis 1 |
| DAB1 | disabled homolog 1 (Drosophila) |
| DAG1 | dystroglycan 1 (dystrophin-associated glycoprotein 1) |
| DAPK1 | death-associated protein kinase 1 |
| DAZL | deleted in azoospermia-like |
| DCAF10 | DDB1 And CUL4 Associated Factor 10 |
| DCAF5 | DDB1 and CUL4 associated factor 5 |
| DCC | deleted in colorectal carcinoma |
| DCHS1 | dachsous 1 (Drosophila) |
| DCP1A | DCP1 decapping enzyme homolog A (S. cerevisiae) |
| DDB1 | damage-specific DNA binding protein 1, 127kDa |
| DDHD1 | DDHD domain containing 1 |
| DDI2 | DNA-damage inducible 1 homolog 2 (S. cerevisiae) |
| DDN | dendrin |
| DDX4 | DEAD (Asp-Glu-Ala-Asp) box polypeptide 4 |
| DDX46 | DEAD (Asp-Glu-Ala-Asp) box polypeptide 46 |
| DENND1A | DENN/MADD domain containing 1A |
| DENND2A | DENN/MADD domain containing 2A |
| DENND4B | DENN/MADD domain containing 4B |
| DENND5A | DENN/MADD domain containing 5A |
| DEPDC5 | DEP domain containing 5 |
| DGKD | diacylglycerol kinase, delta 130kDa |
| DHCR24 | 24-dehydrocholesterol reductase |
| DHX36 | DEAH (Asp-Glu-Ala-His) box polypeptide 36 |
| DHX8 | DEAH (Asp-Glu-Ala-His) box polypeptide 8 |
| DHX9 | DEAH (Asp-Glu-Ala-His) box polypeptide 9 |
| DIDO1 | death inducer-obliterator 1 |
| DIP2B | DIP2 disco-interacting protein 2 homolog B (Drosophila) |
| DKFZP761J1410 | Lipid phosphate phosphatase-related protein type 2 |
| DLC1 | deleted in liver cancer 1 |
| DLG5 | discs, large homolog 5 (Drosophila) |

| | |
|---|---|
| DLGAP2 | discs, large (Drosophila) homolog-associated protein 2 |
| DLGAP3 | discs, large (Drosophila) homolog-associated protein 3 |
| DLGAP4 | discs, large (Drosophila) homolog-associated protein 4 |
| DLL1 | delta-like 1 (Drosophila) |
| DLST | Dihydrolipoamide S-Succinyltransferase |
| DMBX1 | diencephalon/mesencephalon homeobox 1 |
| DMD | dystrophin |
| DMTF1 | cyclin D binding myb-like transcription factor 1 |
| DNAJC2 | DnaJ (Hsp40) homolog, subfamily C, member 2 |
| DNAJC3 | DnaJ (Hsp40) homolog, subfamily C, member 3 |
| DNAJC6 | DnaJ (Hsp40) homolog, subfamily C, member 6 |
| DOCK11 | dedicator of cytokinesis 11 |
| DOCK3 | dedicator of cytokinesis 3 |
| DOCK4 | dedicator of cytokinesis 4 |
| DOCK9 | dedicator of cytokinesis 9 |
| DOPEY1 | dopey family member 1 |
| DPP6 | dipeptidyl-peptidase 6 |
| DPP8 | dipeptidyl-peptidase 8 |
| DPP9 | dipeptidyl-peptidase 9 |
| DRG1 | developmentally regulated GTP binding protein 1 |
| DSCAML1 | Down syndrome cell adhesion molecule like 1 |
| DST | dystonin |
| DVL3 | dishevelled, dsh homolog 3 (Drosophila) |
| DYNC1H1 | dynein, cytoplasmic 1, heavy chain 1 |
| DYNC1LI1 | dynein, cytoplasmic 1, light intermediate chain 1 |
| DYRK1B | dual-specificity tyrosine-(Y)-phosphorylation regulated kinase 1B |
| DYRK2 | dual-specificity tyrosine-(Y)-phosphorylation regulated kinase 2 |
| ECE1 | endothelin converting enzyme 1 |
| EDC4 | enhancer of mRNA decapping 4 |
| EEF2 | eukaryotic translation elongation factor 2 |
| EFEMP1 | EGF containing fibulin-like extracellular matrix protein 1 |
| EFHC2 | EF-hand domain (C-terminal) containing 2 |
| EGFR | epidermal growth factor receptor |
| EGLN1 | egl nine homolog 1 (C. elegans) |

| | |
|---|---|
| EHBP1 | EH domain binding protein 1 |
| EIF2B5 | eukaryotic translation initiation factor 2B, subunit 5 epsilon, 82kDa |
| EIF3D | eukaryotic translation initiation factor 3, subunit D |
| EIF4A2 | eukaryotic translation initiation factor 4A2 |
| EIF4ENIF1 | eukaryotic translation initiation factor 4E nuclear import factor 1 |
| EIF5 | eukaryotic translation initiation factor 5 |
| ELMO1 | engulfment and cell motility 1 |
| EML1 | echinoderm microtubule associated protein like 1 |
| EP400 | E1A binding protein p400 |
| EPHA4 | EPH receptor A4 |
| EPHB1 | EPH receptor B1 |
| EPHB4 | EPH receptor B4 |
| ERBB2 | v-erb-b2 erythroblastic leukemia viral oncogene homolog 2, neuro/glioblastoma derived oncogene homolog (avian) |
| ERBB2IP | erbb2 interacting protein |
| ERBB4 | v-erb-a erythroblastic leukemia viral oncogene homolog 4 (avian) |
| ERC2 | ELKS/RAB6-interacting/CAST family member 2 |
| ESRRG | estrogen-related receptor gamma |
| EYA1 | eyes absent homolog 1 (Drosophila) |
| EYA3 | eyes absent homolog 3 (Drosophila) |
| F2 | coagulation factor II (thrombin) |
| FAM208A | transcription activation suppressor |
| FAM208B | transcription activation suppressor family member 2 |
| FAM214A | atos homolog A |
| FARSB | phenylalanyl-tRNA synthetase, beta subunit |
| FAT3 | FAT tumor suppressor homolog 3 (Drosophila) |
| FAT4 | FAT tumor suppressor homolog 4 (Drosophila) |
| FBRS | fibrosin |
| FBXL20 | F-box and leucine-rich repeat protein 20 |
| FBXO28 | F-box protein 28 |
| FBXO41 | F-box protein 41 |
| FBXO42 | F-box protein 42 |
| FCHO1 | FCH domain only 1 |
| FCHO2 | FCH domain only 2 |

| | |
|---|---|
| FCHSD2 | FCH and double SH3 domains 2 |
| FGD5 | FYVE, RhoGEF and PH domain containing 5 |
| FGF8 | fibroblast growth factor 8 (androgen-induced) |
| FGFR1 | fibroblast growth factor receptor 1 |
| FLCN | folliculin |
| FLNA | filamin A, alpha |
| FLNC | filamin C, gamma |
| FLRT3 | fibronectin leucine rich transmembrane protein 3 |
| FLT1 | fms-related tyrosine kinase 1 (vascular endothelial growth factor/vascular permeability factor receptor) |
| FMN2 | formin 2 |
| FNDC3A | fibronectin type III domain containing 3A |
| FNDC3B | fibronectin type III domain containing 3B |
| FOXJ2 | forkhead box J2 |
| FOXJ3 | forkhead box J3 |
| FOXP1 | forkhead box P1 |
| FRY | furry homolog (Drosophila) |
| FRYL | FRY-like |
| FURIN | furin (paired basic amino acid cleaving enzyme) |
| FUS | fused in sarcoma |
| FXR2 | fragile X mental retardation, autosomal homolog 2 |
| FZD4 | frizzled family receptor 4 |
| G3BP1 | GTPase activating protein (SH3 domain) binding protein 1 |
| G3BP2 | GTPase activating protein (SH3 domain) binding protein 2 |
| GABRA1 | gamma-aminobutyric acid (GABA) A receptor, alpha 1 |
| GABRA5 | gamma-aminobutyric acid (GABA) A receptor, alpha 5 |
| GABRB1 | gamma-aminobutyric acid (GABA) A receptor, beta 1 |
| GABRB2 | gamma-aminobutyric acid (GABA) A receptor, beta 2 |
| GABRG2 | gamma-aminobutyric acid (GABA) A receptor, gamma 2 |
| GANAB | glucosidase, alpha; neutral AB |
| GATAD2A | GATA zinc finger domain containing 2A |
| GCN1L1 | GCN1 general control of amino-acid synthesis 1-like 1 (yeast) |
| GFPT1 | glutamine--fructose-6-phosphate transaminase 1 |
| GIGYF1 | GRB10 interacting GYF protein 1 |

| | |
|---|---|
| GIT1 | G protein-coupled receptor kinase interacting ArfGAP 1 |
| GLI2 | GLI family zinc finger 2 |
| GLTSCR1L | BICRA like chromatin remodeling complex associated protein |
| GLUL | glutamate-ammonia ligase |
| GLYR1 | glyoxylate reductase 1 homolog |
| GMEB2 | glucocorticoid modulatory element binding protein 2 |
| GNAI1 | guanine nucleotide binding protein (G protein), alpha inhibiting activity polypeptide 1 |
| GNAO1 | guanine nucleotide binding protein (G protein), alpha activating activity polypeptide O |
| GNAS | GNAS complex locus |
| GOLGA2 | golgin A2 |
| GOLGA8M | golgin A8 family member M |
| GON4L | gon-4-like (C. elegans) |
| GPR116 | G protein-coupled receptor 116 |
| GPR125 | G protein-coupled receptor 125 |
| GRIA1 | glutamate receptor, ionotropic, AMPA 1 |
| GRIA2 | glutamate receptor, ionotropic, AMPA 2 |
| GRIA4 | glutamate receptor, ionotropic, AMPA 4 |
| GRID2 | glutamate receptor, ionotropic, delta 2 |
| GRIP1 | glutamate receptor interacting protein 1 |
| GRM2 | glutamate receptor, metabotropic 2 |
| GRM3 | glutamate receptor, metabotropic 3 |
| GRM4 | glutamate receptor, metabotropic 4 |
| GRM5 | glutamate receptor, metabotropic 5 |
| GRM7 | glutamate receptor, metabotropic 7 |
| GSPT2 | G1 to S phase transition 2 |
| GTF2E1 | general transcription factor IIE, polypeptide 1, alpha 56kDa |
| GTPBP1 | GTP binding protein 1 |
| HCFC2 | host cell factor C2 |
| HCN1 | hyperpolarization activated cyclic nucleotide-gated potassium channel 1 |
| HDAC5 | histone deacetylase 5 |
| HDAC7 | histone deacetylase 7 |
| HDAC9 | histone deacetylase 9 |
| HDLBP | high density lipoprotein binding protein |

| | |
|---|---|
| HEATR1 | HEAT repeat containing 1 |
| HECTD1 | HECT domain containing E3 ubiquitin protein ligase 1 |
| HECTD4 | HECT domain E3 ubiquitin protein ligase 4 |
| HECW1 | HECT, C2 and WW domain containing E3 ubiquitin protein ligase 1 |
| HERC1 | HECT and RLD domain containing E3 ubiquitin protein ligase family member 1 |
| HERC2 | HECT and RLD domain containing E3 ubiquitin protein ligase 2 |
| HIAT1 | hippocampus abundant transcript 1 |
| HIF1A | hypoxia inducible factor 1, alpha subunit (basic helix-loop-helix transcription factor) |
| HINFP | histone H4 transcription factor |
| HIP1 | huntingtin interacting protein 1 |
| HIRA | HIR histone cell cycle regulation defective homolog A (S. cerevisiae) |
| HIVEP1 | human immunodeficiency virus type I enhancer binding protein 1 |
| HNF4A | hepatocyte nuclear factor 4, alpha |
| HNRNPM | heterogeneous nuclear ribonucleoprotein M |
| HOMER1 | homer homolog 1 (Drosophila) |
| HOXA3 | homeobox A3 |
| HP1BP3 | heterochromatin protein 1, binding protein 3 |
| HSP90B1 | heat shock protein 90kDa beta (Grp94), member 1 |
| HSPA12A | heat shock 70kDa protein 12A |
| HSPA14 | heat shock 70kDa protein 14 |
| HSPA8 | heat shock 70kDa protein 8 |
| HSPD1 | heat shock 60kDa protein 1 (chaperonin) |
| HUNK | hormonally up-regulated Neu-associated kinase |
| HYOU1 | hypoxia up-regulated 1 |
| IGF2R | insulin-like growth factor 2 receptor |
| IGSF3 | immunoglobulin superfamily, member 3 |
| IGSF9B | immunoglobulin superfamily member 9B |
| INCENP | inner centromere protein antigens 135/155kDa |
| INO80D | INO80 complex subunit D |
| INPP4A | inositol polyphosphate-4-phosphatase, type I, 107kDa |
| INTS3 | integrator complex subunit 3 |
| INTS8 | integrator complex subunit 8 |

| | |
|---|---|
| IPO11 | importin 11 |
| IPO8 | importin 8 |
| IQSEC3 | IQ motif and Sec7 domain 3 |
| ITGA5 | integrin, alpha 5 (fibronectin receptor, alpha polypeptide) |
| ITPR1 | inositol 1,4,5-trisphosphate receptor, type 1 |
| ITSN1 | intersectin 1 (SH3 domain protein) |
| IVNS1ABP | influenza virus NS1A binding protein |
| JADE2 | jade family PHD finger 2 |
| JAK3 | Janus kinase 3 |
| JAKMIP1 | janus kinase and microtubule interacting protein 1 |
| JMY | junction mediating and regulatory protein, p53 cofactor |
| JPH3 | junctophilin 3 |
| KALRN | kalirin, RhoGEF kinase |
| KANK2 | KN motif and ankyrin repeat domains 2 |
| KAT2B | K(lysine) acetyltransferase 2B |
| KAT7 | lysine acetyltransferase 7 |
| KCND2 | potassium voltage-gated channel, Shal-related subfamily, member 2 |
| KCNQ4 | potassium voltage-gated channel, KQT-like subfamily, member 4 |
| KCTD3 | potassium channel tetramerisation domain containing 3 |
| KDM3A | lysine (K)-specific demethylase 3A |
| KDM5A | lysine (K)-specific demethylase 5A |
| KDM5C | lysine (K)-specific demethylase 5C |
| KDM7A | lysine demethylase 7A |
| KIAA0232 | KIAA0232 |
| KIAA0907 | KIAA0907 |
| KIAA0922 | KIAA0922 |
| KIAA1244 | KIAA1244 |
| KIAA1429 | KIAA1429 |
| KIAA1549 | KIAA1549 |
| KIF13A | kinesin family member 13A |
| KIF1B | kinesin family member 1B |
| KIF21B | kinesin family member 21B |
| KIF3A | kinesin family member 3A |
| KIF5A | kinesin family member 5A |

| | |
|---|---|
| KIRREL | kin of IRRE like (Drosophila) |
| KIRREL3 | kin of IRRE like 3 (Drosophila) |
| KIT | v-kit Hardy-Zuckerman 4 feline sarcoma viral oncogene homolog |
| KLHL3 | kelch-like 3 (Drosophila) |
| KMT2C | lysine methyltransferase 2C |
| KMT2D | lysine methyltransferase 2D |
| KRT1 | keratin 1 |
| L3MBTL3 | l(3)mbt-like 3 (Drosophila) |
| LAMC1 | laminin, gamma 1 (formerly LAMB2) |
| LARGE | like-glycosyltransferase |
| LCK | lymphocyte-specific protein tyrosine kinase |
| LCP2 | lymphocyte cytosolic protein 2 (SH2 domain containing leukocyte protein of 76kDa) |
| LEMD3 | LEM domain containing 3 |
| LEPR | leptin receptor |
| LHX2 | LIM homeobox 2 |
| LIMCH1 | LIM and calponin homology domains 1 |
| LIMK1 | LIM domain kinase 1 |
| LINGO1 | leucine rich repeat and Ig domain containing 1 |
| LLGL1 | lethal giant larvae homolog 1 (Drosophila) |
| LPHN1 | latrophilin 1 |
| LPHN3 | latrophilin 3 |
| LRCH1 | leucine-rich repeats and calponin homology (CH) domain containing 1 |
| LRP1 | low density lipoprotein receptor-related protein 1 |
| LRP1B | low density lipoprotein receptor-related protein 1B |
| LRP2 | low density lipoprotein receptor-related protein 2 |
| LRP6 | low density lipoprotein receptor-related protein 6 |
| LRP8 | low density lipoprotein receptor-related protein 8, apolipoprotein e receptor |
| LRRC41 | leucine rich repeat containing 41 |
| LRRC4C | leucine rich repeat containing 4C |
| LRRC8A | leucine rich repeat containing 8 family, member A |
| LRRTM2 | leucine rich repeat transmembrane neuronal 2 |
| LTBP3 | latent transforming growth factor beta binding protein 3 |
| LTBP4 | latent transforming growth factor beta binding protein 4 |
| LYST | lysosomal trafficking regulator |

| | |
|---|---|
| MACF1 | microtubule-actin crosslinking factor 1 |
| MAGEL2 | MAGE-like 2 |
| MAGI1 | membrane associated guanylate kinase, WW and PDZ domain containing 1 |
| MAML2 | mastermind-like 2 (Drosophila) |
| MAP1A | microtubule-associated protein 1A |
| MAP2K1 | mitogen-activated protein kinase kinase 1 |
| MAP3K12 | mitogen-activated protein kinase kinase kinase 12 |
| MAP3K4 | mitogen-activated protein kinase kinase kinase 4 |
| MAP3K9 | mitogen-activated protein kinase kinase kinase 9 |
| MAP4 | microtubule-associated protein 4 |
| MAP4K5 | mitogen-activated protein kinase kinase kinase kinase 5 |
| MAP7 | microtubule-associated protein 7 |
| MAPK8IP1 | mitogen-activated protein kinase 8 interacting protein 1 |
| MAPKBP1 | mitogen-activated protein kinase binding protein 1 |
| MAPRE1 | microtubule-associated protein, RP/EB family, member 1 |
| MARK2 | MAP/microtubule affinity-regulating kinase 2 |
| MAST3 | microtubule associated serine/threonine kinase 3 |
| MBD1 | methyl-CpG binding domain protein 1 |
| MBD6 | methyl-CpG binding domain protein 6 |
| MDGA1 | MAM domain containing glycosylphosphatidylinositol anchor 1 |
| MDN1 | MDN1, midasin homolog (yeast) |
| MED1 | mediator complex subunit 1 |
| MED12L | mediator complex subunit 12-like |
| MED13 | mediator complex subunit 13 |
| MED15 | mediator complex subunit 15 |
| MEF2D | myocyte enhancer factor 2D |
| MEGF10 | multiple EGF-like-domains 10 |
| MEGF8 | multiple EGF-like-domains 8 |
| MEPCE | methylphosphate capping enzyme |
| MEST | mesoderm specific transcript homolog (mouse) |
| MET | met proto-oncogene (hepatocyte growth factor receptor) |
| METTL16 | methyltransferase like 16 |
| MFN2 | mitofusin 2 |
| MFSD2A | MFSD2 lysolipid transporter A, lysophospholipid |

| | |
|---|---|
| MICAL3 | microtubule associated monoxygenase, calponin and LIM domain containing 3 |
| MLLT4 | myeloid/lymphoid or mixed-lineage leukemia (trithorax homolog, Drosophila); translocated to, 4 |
| MLLT6 | myeloid/lymphoid or mixed-lineage leukemia (trithorax homolog, Drosophila); translocated to, 6 |
| MLXIP | MLX interacting protein |
| MORC2 | MORC family CW-type zinc finger 2 |
| MORC4 | MORC family CW-type zinc finger 4 |
| MOV10 | Mov10, Moloney leukemia virus 10, homolog (mouse) |
| MPHOSPH8 | M-phase phosphoprotein 8 |
| MPP1 | membrane protein, palmitoylated 1, 55kDa |
| MPPED2 | metallophosphoesterase domain containing 2 |
| MRC2 | mannose receptor, C type 2 |
| MTA2 | metastasis associated 1 family, member 2 |
| MTF1 | metal-regulatory transcription factor 1 |
| MTMR12 | myotubularin related protein 12 |
| MTMR3 | myotubularin related protein 3 |
| MTOR | mechanistic target of rapamycin (serine/threonine kinase) |
| MTPAP | mitochondrial poly(A) polymerase |
| MYCBP2 | MYC binding protein 2, E3 ubiquitin protein ligase |
| MYEF2 | myelin expression factor 2 |
| MYH14 | myosin, heavy chain 14, non-muscle |
| MYO18A | myosin XVIIIA |
| MYO9A | myosin IXA |
| MYO9B | myosin IXB |
| MYT1 | myelin transcription factor 1 |
| NAA15 | N-alpha-acetyltransferase 15, NatA auxiliary subunit |
| NACC1 | nucleus accumbens associated 1, BEN and BTB (POZ) domain containing |
| NAV3 | neuron navigator 3 |
| NCDN | neurochondrin |
| NCOA1 | nuclear receptor coactivator 1 |
| NCOA3 | nuclear receptor coactivator 3 |
| NCOA6 | nuclear receptor coactivator 6 |
| NCOA7 | nuclear receptor coactivator 7 |
| NCOR1 | nuclear receptor corepressor 1 |

| | |
|---|---|
| NCOR2 | nuclear receptor corepressor 2 |
| NDST1 | N-deacetylase/N-sulfotransferase (heparan glucosaminyl) 1 |
| NDUFS2 | NADH dehydrogenase (ubiquinone) Fe-S protein 2, 49kDa (NADH-coenzyme Q reductase) |
| NEMF | nuclear export mediator factor |
| NEURL4 | neuralized homolog 4 (Drosophila) |
| NF1 | neurofibromin 1 |
| NFASC | neurofascin |
| NFATC4 | nuclear factor of activated T-cells, cytoplasmic, calcineurin-dependent 4 |
| NFIA | nuclear factor I/A |
| NFIL3 | nuclear factor, interleukin 3 regulated |
| NFIX | nuclear factor I/X (CCAAT-binding transcription factor) |
| NFKBIZ | nuclear factor of kappa light polypeptide gene enhancer in B-cells inhibitor, zeta |
| NKD1 | naked cuticle homolog 1 (Drosophila) |
| NKTR | natural killer-tumor recognition sequence |
| NLGN2 | neuroligin 2 |
| NMT1 | N-myristoyltransferase 1 |
| NNT | nicotinamide nucleotide transhydrogenase |
| NOP56 | NOP56 ribonucleoprotein homolog (yeast) |
| NOS1 | nitric oxide synthase 1 (neuronal) |
| NPAS2 | neuronal PAS domain protein 2 |
| NR2C2 | nuclear receptor subfamily 2, group C, member 2 |
| NR3C2 | nuclear receptor subfamily 3, group C, member 2 |
| NRXN1 | neurexin 1 |
| NRXN2 | neurexin 2 |
| NRXN3 | neurexin 3 |
| NSD1 | nuclear receptor binding SET domain protein 1 |
| NTN4 | netrin 4 |
| NUMA1 | nuclear mitotic apparatus protein 1 |
| NUP155 | nucleoporin 155kDa |
| NUP188 | nucleoporin 188kDa |
| NUP205 | nucleoporin 205kDa |
| NUP214 | nucleoporin 214kDa |

| | |
|---|---|
| NYAP2 | neuronal tyrosine-phosphorylated phosphoinositide-3-kinase adaptor 2 |
| ODC1 | ornithine decarboxylase 1 |
| ONECUT2 | one cut homeobox 2 |
| OPN5 | opsin 5 |
| OSBP | oxysterol binding protein |
| OSBPL9 | oxysterol binding protein-like 9 |
| OXSR1 | oxidative-stress responsive 1 |
| PABPC1L | poly(A) binding protein, cytoplasmic 1-like |
| PACSIN1 | protein kinase C and casein kinase substrate in neurons 1 |
| PAK7 | p21 protein (Cdc42/Rac)-activated kinase 7 |
| PAN2 | PAN2 poly(A) specific ribonuclease subunit homolog (S. cerevisiae) |
| PAPPA | pregnancy-associated plasma protein A, pappalysin 1 |
| PARK7 | parkinson protein 7 |
| PATL1 | protein associated with topoisomerase II homolog 1 (yeast) |
| PAXBP1 | PAX3 and PAX7 binding protein 1 |
| PBX1 | pre-B-cell leukemia homeobox 1 |
| PCDH17 | protocadherin 17 |
| PCLO | piccolo (presynaptic cytomatrix protein) |
| PCNX | pecanex homolog (Drosophila) |
| PCSK2 | proprotein convertase subtilisin/kexin type 2 |
| PDE2A | phosphodiesterase 2A, cGMP-stimulated |
| PDE4A | phosphodiesterase 4A, cAMP-specific |
| PDGFC | platelet derived growth factor C |
| PDPK1 | 3-phosphoinositide dependent protein kinase-1 |
| PDS5B | PDS5, regulator of cohesion maintenance, homolog B (S. cerevisiae) |
| PDZD2 | PDZ domain containing 2 |
| PDZRN3 | PDZ domain containing ring finger 3 |
| PELI2 | pellino E3 ubiquitin protein ligase family member 2 |
| PELP1 | proline, glutamate and leucine rich protein 1 |
| PHACTR3 | phosphatase and actin regulator 3 |
| PHC3 | polyhomeotic homolog 3 (Drosophila) |
| PHF2 | PHD finger protein 2 |
| PHF20 | PHD finger protein 20 |

| | |
|---|---|
| PHIP | pleckstrin homology domain interacting protein |
| PHKA2 | phosphorylase kinase, alpha 2 (liver) |
| PI4K2A | phosphatidylinositol 4-kinase type 2 alpha |
| PI4KB | phosphatidylinositol 4-kinase, catalytic, beta |
| PICALM | phosphatidylinositol binding clathrin assembly protein |
| PIK3C2B | phosphoinositide-3-kinase, class 2, beta polypeptide |
| PIK3CA | phosphoinositide-3-kinase, catalytic, alpha polypeptide |
| PIK3CB | phosphoinositide-3-kinase, catalytic, beta polypeptide |
| PIK3CD | phosphoinositide-3-kinase, catalytic, delta polypeptide |
| PIK3R2 | phosphoinositide-3-kinase, regulatory subunit 2 (beta) |
| PIKFYVE | phosphoinositide kinase, FYVE finger containing |
| PITPNC1 | phosphatidylinositol transfer protein, cytoplasmic 1 |
| PITPNM2 | phosphatidylinositol transfer protein, membrane-associated 2 |
| PKM | pyruvate kinase M1/2 |
| PLCB3 | phospholipase C, beta 3 (phosphatidylinositol-specific) |
| PLCG2 | phospholipase C, gamma 2 (phosphatidylinositol-specific) |
| PLCL2 | phospholipase C-like 2 |
| PLEKHA5 | pleckstrin homology domain containing, family A member 5 |
| PLEKHA6 | pleckstrin homology domain containing, family A member 6 |
| PLEKHO1 | pleckstrin homology domain containing, family O member 1 |
| PLXNA1 | plexin A1 |
| PLXNA2 | plexin A2 |
| PLXNA4 | plexin A4 |
| PLXND1 | plexin D1 |
| PML | promyelocytic leukemia |
| PNISR | PNN interacting serine and arginine rich protein |
| POLR1A | polymerase (RNA) I polypeptide A, 194kDa |
| POLR2A | polymerase (RNA) II (DNA directed) polypeptide A, 220kDa |
| POLR2B | polymerase (RNA) II (DNA directed) polypeptide B, 140kDa |
| POU2F1 | POU class 2 homeobox 1 |
| PPARGC1A | peroxisome proliferator-activated receptor gamma, coactivator 1 alpha |

| | |
|---|---|
| PPFIA3 | protein tyrosine phosphatase, receptor type, f polypeptide (PTPRF), interacting protein (liprin), alpha 3 |
| PPIG | peptidylprolyl isomerase G (cyclophilin G) |
| PPP1R13B | protein phosphatase 1, regulatory subunit 13B |
| PPP2R3A | protein phosphatase 2, regulatory subunit B", alpha |
| PPP3CB | protein phosphatase 3, catalytic subunit, beta isozyme |
| PPP5C | protein phosphatase 5, catalytic subunit |
| PPP6R1 | protein phosphatase 6 regulatory subunit 1 |
| PPRC1 | peroxisome proliferator-activated receptor gamma, coactivator-related 1 |
| PRDM10 | PR domain containing 10 |
| PRDM2 | PR domain containing 2, with ZNF domain |
| PREX1 | phosphatidylinositol-3,4,5-trisphosphate-dependent Rac exchange factor 1 |
| PRICKLE2 | prickle homolog 2 (Drosophila) |
| PRKACA | protein kinase, cAMP-dependent, catalytic, alpha |
| PRKAR2A | protein kinase, cAMP-dependent, regulatory, type II, alpha |
| PRKCA | protein kinase C, alpha |
| PRKCG | protein kinase C, gamma |
| PRKDC | protein kinase, DNA-activated, catalytic polypeptide |
| PRMT5 | protein arginine methyltransferase 5 |
| PRPF38B | PRP38 pre-mRNA processing factor 38 (yeast) domain containing B |
| PRPF8 | PRP8 pre-mRNA processing factor 8 homolog (S. cerevisiae) |
| PRR12 | proline rich 12 |
| PRR14L | proline rich 14 like |
| PRRC2A | proline rich coiled-coil 2A |
| PRRC2B | proline rich coiled-coil 2B |
| PSIP1 | PC4 and SFRS1 interacting protein 1 |
| PSMD11 | proteasome (prosome, macropain) 26S subunit, non-ATPase, 11 |
| PSME3 | proteasome (prosome, macropain) activator subunit 3 (PA28 gamma; Ki) |
| PSME4 | proteasome (prosome, macropain) activator subunit 4 |
| PTBP3 | polypyrimidine tract binding protein 3 |
| PTCH1 | patched 1 |

| | |
|---|---|
| PTCHD1 | patched domain containing 1 |
| PTK2B | PTK2B protein tyrosine kinase 2 beta |
| PTPN14 | protein tyrosine phosphatase, non-receptor type 14 |
| PTPN2 | protein tyrosine phosphatase, non-receptor type 2 |
| PTPRB | protein tyrosine phosphatase, receptor type, B |
| PTPRK | protein tyrosine phosphatase, receptor type, K |
| PTPRT | protein tyrosine phosphatase, receptor type, T |
| PTPRU | protein tyrosine phosphatase, receptor type, U |
| PUM1 | pumilio homolog 1 (Drosophila) |
| PXK | PX domain containing serine/threonine kinase |
| QRICH1 | glutamine-rich 1 |
| QSER1 | glutamine and serine rich 1 |
| RAB30 | RAB30, member RAS oncogene family |
| RAB3GAP2 | RAB3 GTPase activating protein subunit 2 (non-catalytic) |
| RAB4B | RAB4B, member RAS oncogene family |
| RABGAP1 | RAB GTPase activating protein 1 |
| RAD51 | RAD51 homolog (S. cerevisiae) |
| RAD54L2 | RAD54-like 2 (S. cerevisiae) |
| RAI1 | retinoic acid induced 1 |
| RALGPS2 | Ral GEF with PH domain and SH3 binding motif 2 |
| RANBP2 | RAN binding protein 2 |
| RAP1GAP | RAP1 GTPase activating protein |
| RAP1GAP2 | RAP1 GTPase activating protein 2 |
| RAPGEF5 | Rap guanine nucleotide exchange factor (GEF) 5 |
| RAPH1 | Ras association (RalGDS/AF-6) and pleckstrin homology domains 1 |
| RASGRF2 | Ras protein-specific guanine nucleotide-releasing factor 2 |
| RASGRP2 | RAS guanyl releasing protein 2 (calcium and DAG-regulated) |
| RBBP6 | retinoblastoma binding protein 6 |
| RBL2 | retinoblastoma-like 2 (p130) |
| RBM14 | RNA binding motif protein 14 |
| RBM15 | RNA binding motif protein 15 |
| RBM39 | RNA binding motif protein 39 |
| RBM5 | RNA binding motif protein 5 |
| RC3H1 | ring finger and CCCH-type domains 1 |

| | |
|---|---|
| RC3H2 | ring finger and CCCH-type domains 2 |
| RDX | radixin |
| RELN | reelin |
| REPS1 | RALBP1 associated Eps domain containing 1 |
| RERE | arginine-glutamic acid dipeptide (RE) repeats |
| REV3L | REV3-like, polymerase (DNA directed), zeta, catalytic subunit |
| RFX3 | regulatory factor X, 3 (influences HLA class II expression) |
| RFX4 | regulatory factor X, 4 (influences HLA class II expression) |
| RHOT1 | ras homolog family member T1 |
| RIC8B | resistance to inhibitors of cholinesterase 8 homolog B (C. elegans) |
| RICTOR | RPTOR independent companion of MTOR, complex 2 |
| RIF1 | RAP1 interacting factor homolog (yeast) |
| RMND5A | required for meiotic nuclear division 5 homolog A (S. cerevisiae) |
| RNF123 | ring finger protein 123 |
| RNF220 | ring finger protein 220 |
| RNF40 | ring finger protein 40, E3 ubiquitin protein ligase |
| ROBO2 | roundabout, axon guidance receptor, homolog 2 (Drosophila) |
| ROCK1 | Rho-associated, coiled-coil containing protein kinase 1 |
| ROR1 | receptor tyrosine kinase-like orphan receptor 1 |
| RORC | RAR-related orphan receptor C |
| RPL3 | ribosomal protein L3 |
| RPS3 | ribosomal protein S3 |
| RPS6KA5 | ribosomal protein S6 kinase, 90kDa, polypeptide 5 |
| RSPRY1 | ring finger and SPRY domain containing 1 |
| RTN1 | reticulon 1 |
| RUNDC3A | RUN domain containing 3A |
| RUNX1T1 | runt-related transcription factor 1; translocated to, 1 (cyclin D-related) |
| RXRG | retinoid X receptor, gamma |
| RYR3 | ryanodine receptor 3 |
| SCAF1 | SR-related CTD-associated factor 1 |
| SCAF11 | SR-related CTD-associated factor 11 |

| | |
|---|---|
| SCHIP1 | schwannomin interacting protein 1 |
| SCN2A | sodium channel, voltage-gated, type II, alpha subunit |
| SCN3A | sodium channel, voltage-gated, type III, alpha subunit |
| SCN8A | sodium channel, voltage gated, type VIII, alpha subunit |
| SCRIB | scribbled homolog (Drosophila) |
| SEC24B | SEC24 family, member B (S. cerevisiae) |
| SEMA3A | sema domain, immunoglobulin domain (Ig), short basic domain, secreted, (semaphorin) 3A |
| SEMA3F | sema domain, immunoglobulin domain (Ig), short basic domain, secreted, (semaphorin) 3F |
| SEMA4D | sema domain, immunoglobulin domain (Ig), transmembrane domain (TM) and short cytoplasmic domain, (semaphorin) 4D |
| SEMA6A | sema domain, transmembrane domain (TM), and cytoplasmic domain, (semaphorin) 6A |
| SENP3 | SUMO1/sentrin/SMT3 specific peptidase 3 |
| SEPHS1 | selenophosphate synthetase 1 |
| SETD1A | SET domain containing 1A |
| SETD2 | SET domain containing 2 |
| SF3B3 | splicing factor 3b, subunit 3, 130kDa |
| SFSWAP | splicing factor SWAP |
| SGIP1 | SH3-domain GRB2-like (endophilin) interacting protein 1 |
| SGTB | small glutamine-rich tetratricopeptide repeat (TPR)-containing, beta |
| SHANK1 | SH3 and multiple ankyrin repeat domains 1 |
| SHANK2 | SH3 and multiple ankyrin repeat domains 2 |
| SIK2 | salt-inducible kinase 2 |
| SIN3B | SIN3 transcription regulator homolog B (yeast) |
| SIPA1L1 | signal-induced proliferation-associated 1 like 1 |
| SIPA1L3 | signal-induced proliferation-associated 1 like 3 |
| SIRT1 | sirtuin 1 |
| SKAP2 | src kinase associated phosphoprotein 2 |
| SKI | v-ski sarcoma viral oncogene homolog (avian) |
| SKP2 | S-phase kinase-associated protein 2, E3 ubiquitin protein ligase |
| SLC13A4 | solute carrier family 13 (sodium/sulfate symporters), member 4 |

| | |
|---|---|
| SLC18A2 | solute carrier family 18 (vesicular monoamine), member 2 |
| SLC24A3 | solute carrier family 24 (sodium/potassium/calcium exchanger), member 3 |
| SLC25A19 | solute carrier family 25 (mitochondrial thiamine pyrophosphate carrier), member 19 |
| SLC2A14 | solute carrier family 2 (facilitated glucose transporter), member 14 |
| SLC38A2 | solute carrier family 38, member 2 |
| SLC38A4 | solute carrier family 38, member 4 |
| SLC4A8 | solute carrier family 4, sodium bicarbonate cotransporter, member 8 |
| SLC6A1 | solute carrier family 6 (neurotransmitter transporter, GABA), member 1 |
| SLC6A17 | solute carrier family 6, member 17 |
| SLC6A3 | solute carrier family 6 (neurotransmitter transporter, dopamine), member 3 |
| SLC6A6 | solute carrier family 6 (neurotransmitter transporter, taurine), member 6 |
| SLC7A3 | solute carrier family 7 (cationic amino acid transporter, y+ system), member 3 |
| SLIT2 | slit homolog 2 (Drosophila) |
| SLIT3 | slit homolog 3 (Drosophila) |
| SLITRK1 | SLIT and NTRK-like family, member 1 |
| SLTM | SAFB-like, transcription modulator |
| SMAD5 | SMAD family member 5 |
| SMARCA4 | SWI/SNF related, matrix associated, actin dependent regulator of chromatin, subfamily a, member 4 |
| SMARCAD1 | SWI/SNF-related, matrix-associated actin-dependent regulator of chromatin, subfamily a, containing DEAD/H box 1 |
| SMARCC2 | SWI/SNF related, matrix associated, actin dependent regulator of chromatin, subfamily c, member 2 |
| SMC1B | structural maintenance of chromosomes 1B |
| SMC3 | structural maintenance of chromosomes 3 |
| SMC5 | structural maintenance of chromosomes 5 |
| SMG6 | smg-6 homolog, nonsense mediated mRNA decay factor (C. elegans) |
| SMG7 | smg-7 homolog, nonsense mediated mRNA decay factor (C. elegans) |

| | |
|---|---|
| SMURF1 | SMAD specific E3 ubiquitin protein ligase 1 |
| SNIP1 | Smad nuclear interacting protein 1 |
| SNRNP200 | small nuclear ribonucleoprotein 200kDa (U5) |
| SNW1 | SNW domain containing 1 |
| SNX27 | sorting nexin family member 27 |
| SOGA1 | suppressor of glucose, autophagy associated 1 |
| SORCS1 | sortilin-related VPS10 domain containing receptor 1 |
| SOS1 | son of sevenless homolog 1 (Drosophila) |
| SOX6 | SRY (sex determining region Y)-box 6 |
| SP4 | Sp4 transcription factor |
| SPAST | spastin |
| SPEG | SPEG complex locus |
| SPTB | spectrin, beta, erythrocytic |
| SPTBN1 | spectrin, beta, non-erythrocytic 1 |
| SPTY2D1 | SPT2, Suppressor of Ty, domain containing 1 (S. cerevisiae) |
| SRCIN1 | SRC kinase signaling inhibitor 1 |
| SREBF1 | sterol regulatory element binding transcription factor 1 |
| SREBF2 | sterol regulatory element binding transcription factor 2 |
| SRGAP1 | SLIT-ROBO Rho GTPase activating protein 1 |
| SRGAP3 | SLIT-ROBO Rho GTPase activating protein 3 |
| SRPR | signal recognition particle receptor (docking protein) |
| SRSF6 | serine and arginine rich splicing factor 6 |
| SSH1 | slingshot homolog 1 (Drosophila) |
| STAT1 | signal transducer and activator of transcription 1, 91kDa |
| STAT3 | signal transducer and activator of transcription 3 (acute-phase response factor) |
| STAT5B | signal transducer and activator of transcription 5B |
| STOX2 | storkhead box 2 |
| STRBP | spermatid perinuclear RNA binding protein |
| STXBP1 | syntaxin binding protein 1 |
| STXBP5 | syntaxin binding protein 5 (tomosyn) |
| SUPT16H | suppressor of Ty 16 homolog (S. cerevisiae) |
| SUV420H1 | suppressor of variegation 4-20 homolog 1 (Drosophila) |
| SVIL | supervillin |
| SYDE1 | synapse defective 1, Rho GTPase, homolog 1 (C. elegans) |

| | |
|---|---|
| TAF5L | TAF5-like RNA polymerase II, p300/CBP-associated factor (PCAF)-associated factor, 65kDa |
| TANC2 | tetratricopeptide repeat, ankyrin repeat and coiled-coil containing 2 |
| TBC1D10B | TBC1 domain family, member 10B |
| TBC1D22A | TBC1 domain family, member 22A |
| TCERG1 | transcription elongation regulator 1 |
| TCF12 | transcription factor 12 |
| TCF7L2 | transcription factor 7-like 2 (T-cell specific, HMG-box) |
| TECPR2 | tectonin beta-propeller repeat containing 2 |
| TENM3 | teneurin transmembrane protein 3 |
| TEX10 | testis expressed 10 |
| TEX264 | testis expressed 264 |
| THRB | thyroid hormone receptor, beta |
| THSD7A | thrombospondin, type I, domain containing 7A |
| TJP1 | tight junction protein 1 (zona occludens 1) |
| TLE3 | transducin-like enhancer of split 3 (E(sp1) homolog, Drosophila) |
| TLK2 | tousled-like kinase 2 |
| TLN1 | talin 1 |
| TLN2 | talin 2 |
| TM9SF4 | transmembrane 9 superfamily protein member 4 |
| TMEM108 | transmembrane protein 108 |
| TMEM132D | transmembrane protein 132D |
| TMEM165 | transmembrane protein 165 |
| TMEM63C | transmembrane protein 63C |
| TNIK | TRAF2 and NCK interacting kinase |
| TNPO2 | transportin 2 |
| TNR | tenascin R (restrictin, janusin) |
| TNRC18 | trinucleotide repeat containing 18 |
| TNRC6C | trinucleotide repeat containing 6C |
| TNS3 | tensin 3 |
| TOP1 | topoisomerase (DNA) I |
| TP53BP1 | tumor protein p53 binding protein 1 |
| TP63 | tumor protein p63 |
| TPP2 | tripeptidyl peptidase II |
| TPX2 | TPX2, microtubule-associated, homolog (Xenopus laevis) |

| | |
|---|---|
| TRERF1 | transcriptional regulating factor 1 |
| TRIM33 | tripartite motif containing 33 |
| TRIM8 | tripartite motif containing 8 |
| TRIO | triple functional domain (PTPRF interacting) |
| TRMT12 | tRNA methyltransferase 12 homolog (S. cerevisiae) |
| TRMT1L | tRNA methyltransferase 1 like |
| TRMT5 | tRNA methyltransferase 5 homolog (S. cerevisiae) |
| TRPC4 | transient receptor potential cation channel, subfamily C, member 4 |
| TRPC4AP | transient receptor potential cation channel, subfamily C, member 4 associated protein |
| TRRAP | transformation/transcription domain-associated protein |
| TSHZ1 | teashirt zinc finger homeobox 1 |
| TTBK2 | tau tubulin kinase 2 |
| TTC17 | tetratricopeptide repeat domain 17 |
| TUBB3 | tubulin beta 3 class III |
| TUBGCP3 | tubulin, gamma complex associated protein 3 |
| TULP4 | tubby like protein 4 |
| UBA3 | ubiquitin-like modifier activating enzyme 3 |
| UBA6 | ubiquitin-like modifier activating enzyme 6 |
| UBAP2 | ubiquitin associated protein 2 |
| UBE2Z | ubiquitin-conjugating enzyme E2Z |
| UBE4B | ubiquitination factor E4B |
| UBN1 | ubinuclein 1 |
| UBN2 | ubinuclein 2 |
| UBR2 | ubiquitin protein ligase E3 component n-recognin 2 |
| UBR4 | ubiquitin protein ligase E3 component n-recognin 4 |
| UBTF | upstream binding transcription factor, RNA polymerase I |
| UBXN4 | UBX domain protein 4 |
| UBXN7 | UBX domain protein 7 |
| UCHL1 | ubiquitin carboxyl-terminal esterase L1 (ubiquitin thiolesterase) |
| UHRF1BP1L | UHRF1 binding protein 1-like |
| UHRF2 | ubiquitin-like with PHD and ring finger domains 2, E3 ubiquitin protein ligase |
| UNC79 | unc-79 homolog, NALCN channel complex subunit |
| USP10 | ubiquitin specific peptidase 10 |

| | |
|---|---|
| USP15 | ubiquitin specific peptidase 15 |
| USP19 | ubiquitin specific peptidase 19 |
| USP33 | ubiquitin specific peptidase 33 |
| USP42 | ubiquitin specific peptidase 42 |
| USP47 | ubiquitin specific peptidase 47 |
| USP5 | ubiquitin specific peptidase 5 (isopeptidase T) |
| USP7 | ubiquitin specific peptidase 7 (herpes virus-associated) |
| VCL | vinculin |
| VCP | valosin containing protein |
| VEZF1 | vascular endothelial zinc finger 1 |
| VPS13D | vacuolar protein sorting 13 homolog D (S. cerevisiae) |
| WDFY3 | WD repeat and FYVE domain containing 3 |
| WDR33 | WD repeat domain 33 |
| WDR47 | WD repeat domain 47 |
| WEE1 | WEE1 homolog (S. pombe) |
| WHSC1 | Wolf-Hirschhorn syndrome candidate 1 |
| WNK1 | WNK lysine deficient protein kinase 1 |
| WWP1 | WW domain containing E3 ubiquitin protein ligase 1 |
| XKR6 | XK, Kell blood group complex subunit-related family, member 6 |
| XPO4 | exportin 4 |
| XPO7 | exportin 7 |
| XPOT | exportin, tRNA (nuclear export receptor for tRNAs) |
| XPR1 | xenotropic and polytropic retrovirus receptor 1 |
| XYLT1 | xylosyltransferase I |
| YEATS2 | YEATS domain containing 2 |
| YLPM1 | YLP motif containing 1 |
| ZBTB17 | zinc finger and BTB domain containing 17 |
| ZBTB21 | zinc finger and BTB domain containing 21 |
| ZBTB4 | zinc finger and BTB domain containing 4 |
| ZBTB7C | zinc finger and BTB domain containing 7C |
| ZC3H7B | zinc finger CCCH-type containing 7B |
| ZCCHC6 | zinc finger, CCHC domain containing 6 |
| ZDHHC14 | zinc finger, DHHC-type containing 14 |
| ZEB1 | zinc finger E-box binding homeobox 1 |
| ZFC3H1 | zinc finger, C3H1-type containing |
| ZFHX3 | zinc finger homeobox 3 |

| | |
|---|---|
| ZFHX4 | zinc finger homeobox 4 |
| ZFR | zinc finger RNA binding protein |
| ZFYVE9 | zinc finger, FYVE domain containing 9 |
| ZMYND11 | zinc finger, MYND-type containing 11 |
| ZMYND8 | zinc finger, MYND-type containing 8 |
| ZNF133 | zinc finger protein 133 |
| ZNF407 | zinc finger protein 407 |
| ZNF445 | zinc finger protein 445 |
| ZNF512B | zinc finger protein 512B |
| ZNF521 | zinc finger protein 521 |
| ZNF532 | zinc finger protein 532 |
| ZNF536 | zinc finger protein 536 |
| ZNF574 | zinc finger protein 574 |
| ZNF592 | zinc finger protein 592 |
| ZNF628 | zinc finger protein 628 |
| ZNF629 | zinc finger protein 629 |
| ZNF644 | zinc finger protein 644 |
| ZNF710 | zinc finger protein 710 |
| ZNF711 | zinc finger protein 711 |
| ZNF777 | zinc finger protein 777 |
| ZNF827 | zinc finger protein 827 |
| ZNFX1 | zinc finger, NFX1-type containing 1 |
| ZSWIM8 | zinc finger SWIM-type containing 8 |
| ZZZ3 | zinc finger, ZZ-type containing 3 |
| ZNF606 | zinc finger protein 606 |
| NR1H4 | nuclear receptor subfamily 1, group H, member 4 |
| TUSC2 | tumor suppressor candidate 2 |
| ALX1 | ALX homeobox 1 |
| SNX17 | sorting nexin 17 |
| UBR1 | ubiquitin protein ligase E3 component n-recognin 1 |
| C1orf115 | chromosome 1 open reading frame 115 |
| KCND3 | potassium voltage-gated channel, Shal-related subfamily, member 3 |
| CYB5R3 | cytochrome b5 reductase 3 |
| PRG4 | proteoglycan 4 |
| GPRASP1 | G protein-coupled receptor associated sorting protein 1 |
| IMPAD1 | inositol monophosphatase domain containing 1 |

| | |
|---|---|
| PRKCI | protein kinase C, iota |
| WDR36 | WD repeat domain 36 |
| CNKSR3 | CNKSR family member 3 |
| RFK | riboflavin kinase |
| ST3GAL4 | ST3 beta-galactoside alpha-2,3-sialyltransferase 4 |
| SYNPR | synaptoporin |
| CCL25 | chemokine (C-C motif) ligand 25 |
| ETS1 | v-ets erythroblastosis virus E26 oncogene homolog 1 (avian) |
| SPIN2A | spindlin family, member 2A |
| FUBP3 | far upstream element (FUSE) binding protein 3 |
| OR1C1 | olfactory receptor, family 1, subfamily C, member 1 |
| SYT13 | synaptotagmin XIII |
| TMX2 | thioredoxin-related transmembrane protein 2 |
| OR7G3 | olfactory receptor, family 7, subfamily G, member 3 |
| CASS4 | Cas scaffolding protein family member 4 |
| DYNLRB2 | dynein, light chain, roadblock-type 2 |
| TSKU | tsukushi small leucine rich proteoglycan homolog (Xenopus laevis) |
| GPR158 | G protein-coupled receptor 158 |
| HSD17B6 | hydroxysteroid (17-beta) dehydrogenase 6 homolog (mouse) |
| SEC14L1 | SEC14-like 1 (S. cerevisiae) |
| ADIPOR1 | adiponectin receptor 1 |
| CBX4 | chromobox homolog 4 |
| FAM81A | family with sequence similarity 81, member A |
| GRAP2 | GRB2-related adaptor protein 2 |
| KCNK5 | potassium channel, subfamily K, member 5 |
| KRTAP17-1 | keratin associated protein 17-1 |
| LIN28A | lin-28 homolog A |
| NMT2 | N-myristoyltransferase 2 |
| PPP2R1A | protein phosphatase 2, regulatory subunit A, alpha |
| RAB24 | RAB24, member RAS oncogene family |
| REEP2 | receptor accessory protein 2 |
| SERPINB5 | serpin peptidase inhibitor, clade B (ovalbumin), member 5 |
| ZBTB7B | zinc finger and BTB domain containing 7B |
| HOMEZ | homeobox and leucine zipper encoding |

| | |
|---|---|
| RNASE1 | ribonuclease, RNase A family, 1 (pancreatic) |
| SMOX | spermine oxidase |
| ST8SIA2 | ST8 alpha-N-acetyl-neuraminide alpha-2,8-sialyltransferase 2 |
| ATF7 | activating transcription factor 7 |
| KCNB1 | potassium voltage-gated channel, Shab-related subfamily, member 1 |
| PAX5 | paired box 5 |
| CHD2 | chromodomain helicase DNA binding protein 2 |
| HNRNPU | heterogeneous nuclear ribonucleoprotein U (scaffold attachment factor A) |
| ZMYM2 | zinc finger, MYM-type 2 |
| BMPR1B | bone morphogenetic protein receptor, type IB |
| KCMF1 | potassium channel modulatory factor 1 |
| KDM5B | lysine (K)-specific demethylase 5B |
| YPEL4 | yippee-like 4 (Drosophila) |
| CCAR2 | cell cycle and apoptosis regulator 2 |
| AK4 | adenylate kinase 4 |
| FGF9 | fibroblast growth factor 9 (glia-activating factor) |
| GPR34 | G protein-coupled receptor 34 |
| CYTH2 | cytohesin 2 |
| GPHN | gephyrin |
| SMAD2 | SMAD family member 2 |
| CDK5 | cyclin-dependent kinase 5 |
| SMC1A | structural maintenance of chromosomes 1A |
| KCNJ6 | potassium inwardly-rectifying channel, subfamily J, member 6 |
| LTBR | lymphotoxin beta receptor (TNFR superfamily, member 3) |
| MIA3 | melanoma inhibitory activity family, member 3 |
| SRP9 | signal recognition particle 9kDa |
| HNF1B | HNF1 homeobox B |
| GAPDH | glyceraldehyde-3-phosphate dehydrogenase |
| KLHL26 | kelch-like 26 (Drosophila) |
| MADD | MAP-kinase activating death domain |
| RPL38 | ribosomal protein L38 |
| TMEM178A | transmembrane protein 178A |
| BCKDK | branched chain ketoacid dehydrogenase kinase |

| | |
|---|---|
| UBE2Q1 | ubiquitin-conjugating enzyme E2Q family member 1 |
| HIST3H2A | histone cluster 3, H2a |
| LNX2 | ligand of numb-protein X 2 |
| CLEC16A | C-type lectin domain family 16, member A |
| DCAF12L1 | DDB1 and CUL4 associated factor 12 like 1 |
| PDZD8 | PDZ domain containing 8 |
| RAB6A | RAB6A, member RAS oncogene family |
| SV2A | synaptic vesicle glycoprotein 2A |
| MMP16 | matrix metallopeptidase 16 (membrane-inserted) |
| STT3A | STT3, subunit of the oligosaccharyltransferase complex, homolog A (S. cerevisiae) |
| ATP6V0D1 | ATPase, H+ transporting, lysosomal 38kDa, V0 subunit d1 |
| FZD7 | frizzled family receptor 7 |
| FRG2 | FSHD region gene 2 |
| KCNS2 | potassium voltage-gated channel, delayed-rectifier, subfamily S, member 2 |
| EFTUD2 | elongation factor Tu GTP binding domain containing 2 |
| MED12 | mediator complex subunit 12 |
| ERLIN1 | ER lipid raft associated 1 |
| GNB1 | guanine nucleotide binding protein (G protein), beta polypeptide 1 |
| PICK1 | protein interacting with PRKCA 1 |
| ADAMTS10 | ADAM metallopeptidase with thrombospondin type 1 motif, 10 |
| SAMD8 | sterile alpha motif domain containing 8 |
| HIST2H2AB | histone cluster 2, H2ab |
| HDAC3 | histone deacetylase 3 |
| KLHL31 | kelch-like 31 (Drosophila) |
| SOX17 | SRY (sex determining region Y)-box 17 |
| MED23 | mediator complex subunit 23 |
| CRAMP1L | Crm, cramped-like (Drosophila) |
| EIF2S1 | eukaryotic translation initiation factor 2, subunit 1 alpha, 35kDa |
| FBXL3 | F-box and leucine-rich repeat protein 3 |
| RHOA | ras homolog family member A |
| SPCS2 | signal peptidase complex subunit 2 homolog (S. cerevisiae) |

| | |
|---|---|
| TOMM22 | translocase of outer mitochondrial membrane 22 homolog (yeast) |
| CSNK2A2 | casein kinase 2, alpha prime polypeptide |
| ZYG11B | zyg-11 homolog B (C. elegans) |
| NPR2 | natriuretic peptide receptor B/guanylate cyclase B (atrionatriuretic peptide receptor B) |

*This table lists *n*=1046 genes used to perform enrichment analyses with iFC maps in the present study. The list combines two sets of genes (set 1 n=932, set 2 n=114) selected from a larger list of genes analyzed in Satterstrom et al (2019). Set 1 encompasses *n*=932 genes with similarly high rate of protein truncating variants (PTVs) in ASD and ADHD including genes with high probability (>0.9) of loss-of-function intolerant (pLI) (see the original supplementary file 3 in Satterstrom et al., 2019). Set 2 includes *n*=114 genes with both PTVs at pLI<0.9 (without including the 932 mentioned above) and Missense (Mis) variants at pLI above and below 0.9 that we identified based on two-tailed Fisher z exact tests comparing the rate of variants between 'cases' (i.e., ASD and ADHD) without intellectual disability vs. controls among 1,514 genes listed in Satterstrom et al's original supplementary file 4 (Satterstrom et al. 2019).

**Table S5.** Comorbid psychiatric diagnoses information in the primary diagnositc groups

| Variable | Combined Sample | ASD | $ADHD_{w/o\ ASD}$ |
|---|---|---|---|
| | N=166 | n=63 | n=103 |
| **Number of psychiatric comorbid diagnoses** #, (%) | | | |
| No Comorbidities | 58 (35) | 4 (6) | 54 (52) |
| 1 Comorbidity | 58 (35) | 25 (40) | 33 (32) |
| > 1 Comorbidities | 50 (30) | 34 (54) | 16 (16) |
| **Type of Comorbid disorders,** #, (%) | | | |
| ADHD only | NA | 22 (35) | NA |
| Any Anxiety disorder only[a] | 21 (13) | 3 (5) | 18 (17) |
| Behavioral Disruptive Disorders only[b] | 10 (6) | 1 (2) | 9 (9) |
| ADHD + Anxiety[c] | 18 (11) | 18 (29) | NA |
| ADHD + other behavioral disruptive disorders[d] | 5 (3) | 5 (8) | NA |
| Anxiety + Mood disorders[e] | 6 (4) | 1 (2) | 5 (5) |
| SCD only and combined with others [f] | 2 (1) | NA | 2 (2) |
| Elimination only and combined with other disorders [g] | 16 (10) | 8 (13) | 8 (8) |
| Specific Learning Disorders only and combined with others [h] | 6 (4) | 1* (2) | 5 (5) |
| Tic disorders only or combined with others[i] | 3 (2) | 1 (2) | 2 (2) |

**a. n**=16 children with ADHDw/oASD had a comorbid anxiety disorder alone: anxiety NOS (5), GAD (3), separation anxiety (3), social phobia (2) and specific phobia (3). And 2 had 2 comorbid anxiety disorders: social phobia in combination with specific phobia (1) and separation anxiety (1). 2 children with ASD had a comorbid anxiety disorder alone : anxiety NOS (1) and social phobia (1). 1 child with ASD had comorbid separation anxiety and GAD;**b.** 8 children with ADHD and 1 child with ASD had comorbid ODD only. 1 child with ADHD had a comorbid other disruptive impulse-control disorder;**c.** 18 children with ASD with comorbid ADHD in combination with one anxiety disorder (13): specific phobia (5), anxiety NOS (3), GAD (2), social phobia (1) and separation anxiety (2); 1 children combined with ODD and separation anxiety and 4 combined with specific phobia (1 with ODD, 1 with anxiety NOS, 1 with anxiety NOS and mood NOS and 1 with social phobia, GAD and separation anxiety);**d.** 5 children with ASD and comorbid ADHD combined with ODD (from which 1 child had a second disruptive control disorder);**e.** 3 children with ADHD and comorbid mood NOS combined with anxiety NOS (1), with OCD (1, also combined with DMDD) and with GAD and specific learning disorder (1). 1 children with ADHD and comorbid ODD combined with GAD.1 child with ADHD and comorbid dysthimia with separation anxiety. 1 child with ASD and comorbid specific phobia in combination with DMDD;**f.** 2 children with ADHD : one with comorbid SCD alone and one in combination with social phobia and GAD**g.** From those 8 children with ADHD and comorbid elimination disorders, 4 had comorbid enuresis only and 2 had comorbid enuresis combined with GAD. 2 children had both enuresis and encopresis combined with ODD (1) and specific language disorder (1). From those 8 with ASD, 2 comorbid encopresis and 4 enuresis combined with ADHD; and 1 child comorbid enureis and encopresis in combination with ADHD and ODD. 1 child with ASD had comorbid enuresis with specific learning disorder;**i.** From those 5 children with ADHD with comorbid specific learning disorders, 3 were alone and 2 combined (2 with anxiety NOS, from which 1 with OCD and ODD). A child with ASD had comorbid specific learning disorder combined with enuresis (double counted*)**j.** From those 2 children with ADHD with comorbid tic disorder, one child had comorbid tic disorder alone and 1 combined with ODD and a third comorbid encopresis disorder. 1 child with ASD had a comorbid tic disorder combined with ADHD.

**Table S6.** List of the $n$=107 genes enriched among those expressed across middle frontal gyrus iFC map.

| Gene symbol | Gene name |
|---|---|
| ABR | active BCR-related |
| ACTN2 | actinin, alpha 2 |
| AP2S1 | adaptor-related protein complex 2, sigma 1 subunit |
| ARHGEF17 | Rho guanine nucleotide exchange factor (GEF) 17 |
| ARID5B | AT rich interactive domain 5B (MRF1-like) |
| ASTN1 | astrotactin 1 |
| ASXL3 | additional sex combs like 3 (Drosophila) |
| BAI2 | brain-specific angiogenesis inhibitor 2 |
| BTBD11 | BTB (POZ) domain containing 11 |
| C20orf112 | chromosome 20 open reading frame 112 |
| CACNB1 | calcium channel, voltage-dependent, beta 1 subunit |
| CACNG3 | calcium channel, voltage-dependent, gamma subunit 3 |
| CDH9 | cadherin 9, type 2 (T1-cadherin) |
| CHRM3 | cholinergic receptor, muscarinic 3 |
| COL12A1 | collagen, type XII, alpha 1 |
| CSNK2A2 | casein kinase 2, alpha prime polypeptide |
| CTNNA2 | catenin (cadherin-associated protein), alpha 2 |
| CTNND2 | catenin (cadherin-associated protein), delta 2 (neural plakophilin-related arm-repeat protein) |
| CUX2 | cut-like homeobox 2 |
| DIDO1 | death inducer-obliterator 1 |
| DLGAP2 | discs, large (Drosophila) homolog-associated protein 2 |
| DMBX1 | diencephalon/mesencephalon homeobox 1 |
| DOCK3 | dedicator of cytokinesis 3 |
| DPP6 | dipeptidyl-peptidase 6 |
| ECE1 | endothelin converting enzyme 1 |
| EPHB1 | EPH receptor B1 |
| EPHB4 | EPH receptor B4 |
| ERBB4 | v-erb-a erythroblastic leukemia viral oncogene homolog 4 (avian) |
| ERC2 | ELKS/RAB6-interacting/CAST family member 2 |
| FAM81A | family with sequence similarity 81, member A |
| FGD5 | FYVE, RhoGEF and PH domain containing 5 |
| FMN2 | formin 2 |
| FZD4 | frizzled family receptor 4 |
| FZD7 | frizzled family receptor 7 |
| G3BP2 | GTPase activating protein (SH3 domain) binding protein 2 |
| GABRG2 | gamma-aminobutyric acid (GABA) A receptor, gamma 2 |
| GPRASP1 | G protein-coupled receptor associated sorting protein 1 |
| GRM3 | glutamate receptor, metabotropic 3 |
| HIST3H2A | histone cluster 3, H2a |
| HIVEP1 | human immunodeficiency virus type I enhancer binding protein 1 |
| HSPA12A | heat shock 70kDa protein 12A |
| IGSF3 | immunoglobulin superfamily, member 3 |
| JAKMIP1 | janus kinase and microtubule interacting protein 1 |
| KCNB1 | potassium voltage-gated channel, Shab-related subfamily, member 1 |
| KCND2 | potassium voltage-gated channel, Shal-related subfamily, member 2 |
| KCNS2 | potassium voltage-gated channel, delayed-rectifier, subfamily S, member 2 |
| KIF5A | kinesin family member 5A |
| KIT | v-kit Hardy-Zuckerman 4 feline sarcoma viral oncogene homolog |
| LCP2 | lymphocyte cytosolic protein 2 (SH2 domain containing leukocyte protein of 76kDa) |
| LHX2 | LIM homeobox 2 |
| LIMK1 | LIM domain kinase 1 |
| LRCH1 | leucine-rich repeats and calponin homology (CH) domain containing 1 |
| LRRC4C | leucine rich repeat containing 4C |
| MAML2 | mastermind-like 2 (Drosophila) |
| MAP1A | microtubule-associated protein 1A |
| MAST3 | microtubule associated serine/threonine kinase 3 |
| MED13 | mediator complex subunit 13 |
| MED15 | mediator complex subunit 15 |
| MMP16 | matrix metallopeptidase 16 (membrane-inserted) |
| MYT1 | myelin transcription factor 1 |
| NFIA | nuclear factor I/A |
| NPR2 | natriuretic peptide receptor B/guanylate cyclase B (atrionatriuretic peptide receptor B) |
| NTN4 | netrin 4 |
| NUP205 | nucleoporin 205kDa |
| PATL1 | protein associated with topoisomerase II homolog 1 (yeast) |
| PBX1 | pre-B-cell leukemia homeobox 1 |
| PCSK2 | proprotein convertase subtilisin/kexin type 2 |
| PDE2A | phosphodiesterase 2A, cGMP-stimulated |
| PDZD2 | PDZ domain containing 2 |
| PDZRN3 | PDZ domain containing ring finger 3 |
| PELI2 | pellino E3 ubiquitin protein ligase family member 2 |
| PHC3 | polyhomeotic homolog 3 (Drosophila) |
| PLCB3 | phospholipase C, beta 3 (phosphatidylinositol-specific) |
| PLXND1 | plexin D1 |
| PRPF38B | PRP38 pre-mRNA processing factor 38 (yeast) domain containing B |
| PTPRK | protein tyrosine phosphatase, receptor type, K |
| PTPRT | protein tyrosine phosphatase, receptor type, T |
| RASGRF2 | Ras protein-specific guanine nucleotide-releasing factor 2 |
| REV3L | REV3-like, polymerase (DNA directed), zeta, catalytic subunit |
| SCAF11 | SR-related CTD-associated factor 11 |
| SEC24B | SEC24 family, member B (S. cerevisiae) |
| SEMA3A | sema domain, immunoglobulin domain (Ig), short basic domain, secreted, (semaphorin) 3A |
| SGTB | small glutamine-rich tetratricopeptide repeat (TPR)-containing, beta |
| SHANK1 | SH3 and multiple ankyrin repeat domains 1 |
| SIK2 | salt-inducible kinase 2 |
| SIPA1L3 | signal-induced proliferation-associated 1 like 3 |
| SIRT1 | sirtuin 1 |
| SKAP2 | src kinase associated phosphoprotein 2 |
| SKP2 | S-phase kinase-associated protein 2, E3 ubiquitin protein ligase |
| SLC24A3 | solute carrier family 24 (sodium/potassium/calcium exchanger), member 3 |
| SLIT2 | slit homolog 2 (Drosophila) |
| SLITRK1 | SLIT and NTRK-like family, member 1 |
| SORCS1 | sortilin-related VPS10 domain containing receptor 1 |
| SOX6 | SRY (sex determining region Y)-box 6 |
| TAF5L | TAF5-like RNA polymerase II, p300/CBP-associated factor (PCAF)-associated factor, 65kDa |
| TANC2 | tetratricopeptide repeat, ankyrin repeat and coiled-coil containing 2 |
| TCERG1 | transcription elongation regulator 1 |
| TMEM132D | transmembrane protein 132D |
| TPP2 | tripeptidyl peptidase II |
| TSKU | tsukushi small leucine rich proteoglycan homolog (Xenopus laevis) |
| TTC17 | tetratricopeptide repeat domain 17 |
| UBN2 | ubinuclein 2 |
| UCHL1 | ubiquitin carboxyl-terminal esterase L1 (ubiquitin thiolesterase) |
| USP5 | ubiquitin specific peptidase 5 (isopeptidase T) |
| ZDHHC14 | zinc finger, DHHC-type containing 14 |
| ZNF521 | zinc finger protein 521 |
| ZNF532 | zinc finger protein 532 |

**Table S7.** Genetic Ontology (GO) terms significantly associated with the n=107 genes enriched in our MFG iFC map sorted following their hierarchical relations provided by the GO database.

| GO Biological Process | # genes | Expected | Fold enrichment | FDR-p value |
|---|---|---|---|---|
| axonogenesis / axonogenesis involved in innervation | 14/3 | 1.9/0.05 | 7.35/64.75 | **0.0000094**/0.00488 |
| cell morphogenesis involved in neuron differentiation | 17 | 2.31 | 7.36 | **0.000002** |
| neuron development | 20 | 4.47 | 4.47 | **0.000018** |
| cell development | 30 | 11.53 | 2.6 | 0.000408 |
| cell differentiation | 36 | 18.81 | 1.91 | 0.016100 |
| cellular developmental process | 36 | 18.82 | 1.91 | 0.016000 |
| developmental process | 50 | 29.54 | 1.69 | **0.009070** |
| anatomical structure development | 49 | 26.93 | 1.82 | 0.002760 |
| neuron differentiation | 25 | 5.64 | 4.44 | **0.000001** |
| generation of neurons | 26 | 6.03 | 4.31 | **0.000001** |
| neurogenesis | 27 | 6.93 | 3.9 | **0.000002** |
| nervous system development | 35 | 11.36 | 3.08 | **0.000002** |
| system development | 44 | 18.25 | 2.41 | **0.000009** |
| multicellular organism development | 46 | 20.41 | 2.25 | **0.000018** |
| multicellular organismal process | 57 | 34.72 | 1.64 | 0.004380 |
| cell morphogenesis | 20 | 3.58 | 5.58 | **0.000001** |
| anatomical structure morphogenesis | 32 | 11.53 | 2.78 | **0.000042** |
| neuron projection morphogenesis | 16 | 2.53 | 6.33 | **0.000010** |
| neuron projection development | 17 | 3.61 | 4.7 | **0.000091** |
| plasma membrane bounded cell projection organization | 21 | 5.95 | 3.53 | **0.000284** |
| cell projection organization | 21 | 6.2 | 3.39 | 0.000486 |
| cellular component organization | 48 | 29.01 | 1.65 | 0.019400 |
| cellular component organization or biogenesis | 48 | 30.12 | 1.59 | 0.041700 |
| plasma membrane bounded cell projection morphogenesis | 16 | 2.55 | 6.27 | **0.000010** |
| cell projection morphogenesis | 16 | 2.58 | 6.2 | **0.000009** |
| cellular anatomical entity morphogenesis | 17 | 3.15 | 5.4 | **0.000017** |
| axon development | 14 | 2.18 | 6.43 | **0.000035** |
| dendritic spine morphogenesis | 3 | 0.09 | 32.38 | 0.024000 |
| neuron projection organization | 4 | 0.23 | 17.66 | 0.020200 |
| synapse organization | 13 | 1.77 | 7.36 | **0.000024** |
| cell junction organization | 15 | 2.78 | 5.39 | **0.000093** |
| dendrite development | 6 | 0.59 | 10.22 | 0.008900 |
| dendrite morphogenesis | 5 | 0.33 | 15.18 | 0.007140 |
| olfactory bulb development | 4 | 0.18 | 22.2 | 0.009550 |
| olfactory lobe development | 4 | 0.19 | 21 | 0.011500 |
| brain development | 13 | 3.82 | 3.4 | 0.025700 |
| head development | 14 | 4.08 | 3.43 | 0.016100 |
| axon extension | 4 | 0.24 | 16.53 | 0.024900 |
| developmental growth | 11 | 2.22 | 4.95 | 0.005690 |
| growth | 11 | 2.22 | 4.95 | 0.005840 |
| ovulation cycle process | 4 | 0.24 | 16.53 | 0.024500 |
| protein localization to synapse | 5 | 0.32 | 15.42 | 0.006770 |
| protein localization to cell junction | 5 | 0.46 | 10.79 | 0.024300 |
| female gonad development | 6 | 0.48 | 12.53 | 0.004070 |
| development of primary female sexual characteristics | 6 | 0.5 | 12.02 | 0.004750 |
| female sex differentiation | 6 | 0.58 | 10.41 | 0.008600 |
| regulation of axonogenesis | 6 | 0.81 | 7.38 | 0.035600 |
| regulation of neuron projection development | 11 | 2.36 | 4.66 | 0.008410 |
| regulation of plasma membrane bounded cell projection organization | 12 | 3.38 | 3.55 | 0.031300 |
| regulation of cell projection organization | 12 | 3.46 | 3.47 | 0.036400 |
| regulation of cellular process | 79 | 57.48 | 1.37 | 0.007430 |
| regulation of biological process | 85 | 60.34 | 1.41 | 0.000407 |
| biological regulation | 86 | 62.5 | 1.38 | 0.000818 |
| regulation of anatomical structure morphogenesis | 15 | 4.35 | 3.45 | 0.008800 |
| ameboidal-type cell migration | 7 | 1.03 | 6.8 | 0.020500 |
| axon guidance | 7 | 1.17 | 5.96 | 0.036400 |
| neuron projection guidance | 7 | 1.17 | 5.96 | 0.035900 |
| positive regulation of nervous system development | 8 | 1.51 | 5.29 | 0.031600 |
| regulation of nervous system development | 10 | 2.39 | 4.19 | 0.030800 |
| modulation of chemical synaptic transmission | 13 | 2.48 | 5.24 | 0.000752 |
| regulation of trans-synaptic signaling | 13 | 2.49 | 5.23 | 0.000738 |
| regulation of cell communication | 34 | 17.71 | 1.92 | 0.025500 |
| regulation of signaling | 35 | 17.68 | 1.98 | 0.010700 |
| chemical synaptic transmission | 10 | 2.16 | 4.63 | 0.017300 |
| anterograde trans-synaptic signaling | 10 | 2.16 | 4.63 | 0.017000 |
| trans-synaptic signaling | 10 | 2.25 | 4.44 | 0.022400 |
| synaptic signaling | 10 | 2.4 | 4.16 | 0.032000 |
| signaling | 47 | 26.3 | 1.79 | 0.005370 |
| cell communication | 49 | 27.09 | 1.81 | 0.002780 |
| regulation of neurogenesis | 9 | 2 | 4.49 | 0.036200 |
| enzyme-linked receptor protein signaling pathway | 13 | 3.16 | 4.11 | 0.006200 |
| cell surface receptor signaling pathway | 26 | 10.7 | 2.43 | 0.005570 |
| behavior | 12 | 3.25 | 3.69 | 0.024200 |
| cell adhesion | 17 | 4.92 | 3.45 | 0.003780 |
| animal organ morphogenesis | 15 | 5.13 | 2.93 | 0.036600 |
| regulation of molecular function | 26 | 10.34 | 2.52 | 0.003770 |
| regulation of biological quality | 33 | 14.67 | 2.25 | 0.003150 |

*Only genetic ontology (GO) terms surviving FDR correction at q<0.05 are shown. Gene ontology database sorts biological process terms based on their hierarchical relations going from the most specific to more general terms. Multiple terms may share hierarchical relationships with more general terms in that case the "parent" term would appear indented directly below the specific term. Examples of broad biological process terms are *mullticellular organismal processe*s or *nervous system development*. Examples of more specific terms are *axonogenesis* or *dendritic spine morphogenesis*. The top 20 statistically signifcant terms are indicated as bold text. Most these terms are associated to the parent term *neuron projection morphogenesis* and their most specific related terms are *axonogeneis* and *regulation of axonogenesis*, *dendritic spine morphogenesis*, *axon extension* and *axon guidance*, their hierarchical relationship and definitions are described in Figure S6.
